# Interventions targeting nonsymptomatic cases can be important to prevent local outbreaks: SARS-CoV-2 as a case-study

**DOI:** 10.1101/2020.11.06.20226969

**Authors:** Francesca A. Lovell-Read, Sebastian Funk, Uri Obolski, Christl A. Donnelly, Robin N. Thompson

**Author notes:** **Corresponding author** Address: Merton College, Merton Street, Oxford, OX1 4JD.

## Abstract

During infectious disease epidemics, an important question is whether cases travelling to new locations will trigger local outbreaks. The risk of this occurring depends on the transmissibility of the pathogen, the susceptibility of the host population and, crucially, the effectiveness of surveillance in detecting cases and preventing onward spread. For many pathogens, transmission from presymptomatic and/or asymptomatic (together referred to as nonsymptomatic) infectious hosts can occur, making effective surveillance challenging. Here, using SARS-CoV-2 as a case-study, we show how the risk of local outbreaks can be assessed when nonsymptomatic transmission can occur. We construct a branching process model that includes nonsymptomatic transmission, and explore the effects of interventions targeting nonsymptomatic or symptomatic hosts when surveillance resources are limited. We consider whether the greatest reductions in local outbreak risks are achieved by increasing surveillance and control targeting nonsymptomatic or symptomatic cases, or a combination of both. We find that seeking to increase surveillance of symptomatic hosts alone is typically not the optimal strategy for reducing outbreak risks. Adopting a strategy that combines an enhancement of surveillance of symptomatic cases with efforts to find and isolate nonsymptomatic infected hosts leads to the largest reduction in the probability that imported cases will initiate a local outbreak.

## 1. Introduction

Emerging epidemics represent a substantial challenge to human health worldwide [1-4]. When cases are clustered in specific locations, two key questions are: i) Will exported cases lead to local outbreaks in new locations? and ii) Which surveillance and control strategies in those new locations will reduce the risk of local outbreaks?

Branching process models are used for a range of diseases to assess whether cases that are newly arrived in a host population will generate a local outbreak driven by sustained local transmission [5-11]. These models can also be used to predict the effectiveness of potential control interventions. For example, early in the COVID-19 pandemic, Hellewell *et al*. [12] used simulations of a branching process model to predict whether or not new outbreaks would fade out under different contact tracing strategies. Thompson [13] estimated the probability of local outbreaks analytically using a branching process model and found that effective isolation of infectious hosts leads to a substantial reduction in the outbreak risk.

A factor that can hinder control interventions during any epidemic is the potential for individuals to transmit a pathogen while not showing symptoms. For COVID-19, the incubation period has been estimated to last approximately five or six days on average [14, 15], and presymptomatic transmission can occur during that period [16-20]. Additionally, asymptomatic infected individuals (those who never develop symptoms) are also thought to contribute to transmission [16, 21, 22].

Motivated by the need to assess the risk of outbreaks outside China early in the COVID-19 pandemic, we show how the risk that imported cases will lead to local outbreaks can be estimated using a branching process model. Unlike standard approaches for estimating the probability of a major epidemic analytically [23-26], nonsymptomatic individuals are included in the model explicitly. Using a function that characterises the efficacy of interventions for different surveillance efforts (denoted *f* (*ρ, δ*) in the model), we explore the effects of interventions that aim to reduce this risk. Under the assumption that detected infected hosts are isolated effectively, we consider whether it is most effective to dedicate resources to enhancing surveillance targeting symptomatic individuals, to instead focus on increasing surveillance for nonsymptomatic individuals, or to use a combination of these approaches.

We show that, when surveillance resources are limited, the maximum reduction in the outbreak risk almost always corresponds to a mixed strategy involving enhanced surveillance of both symptomatic and nonsymptomatic hosts. This remains the case even if the surveillance effort required to find nonsymptomatic infected individuals is significantly larger than the effort required to find symptomatic individuals. This highlights the benefits of not only seeking to find and isolate symptomatic hosts, but also dedicating resources to detecting nonsymptomatic cases during infectious disease epidemics.

## 2. Methods

### 2.1 Model

We consider a branching process model in which infectious individuals are classified as asymptomatic (*A*), presymptomatic (*I*_1_) or symptomatic (*I*_2_). Hosts in any of these classes may generate new infections. The parameter *ξ* represents the proportion of new infections that are asymptomatic, so that a new infection either involves increasing *A* by one (with probability *ξ*) or increasing *I*_1_ by one (with probability 1 − *ξ*).

Presymptomatic hosts may go on to develop symptoms (transition from *I*_1_ to *I*_2_) or be detected and isolated (so that *I*_1_ decreases by one). Symptomatic individuals (*I*_2_) can be isolated (so that *I*_2_ decreases by one) or be removed due to recovery or death (so that again *I*_2_ decreases by one). Similarly, asymptomatic hosts may be detected and isolated, or recover (so that *A* decreases by one in either case).

A schematic showing the different possible events in the model is shown in Fig 1A. The analogous compartmental differential equation model to the branching process model that we consider is given by

**Fig 1.**
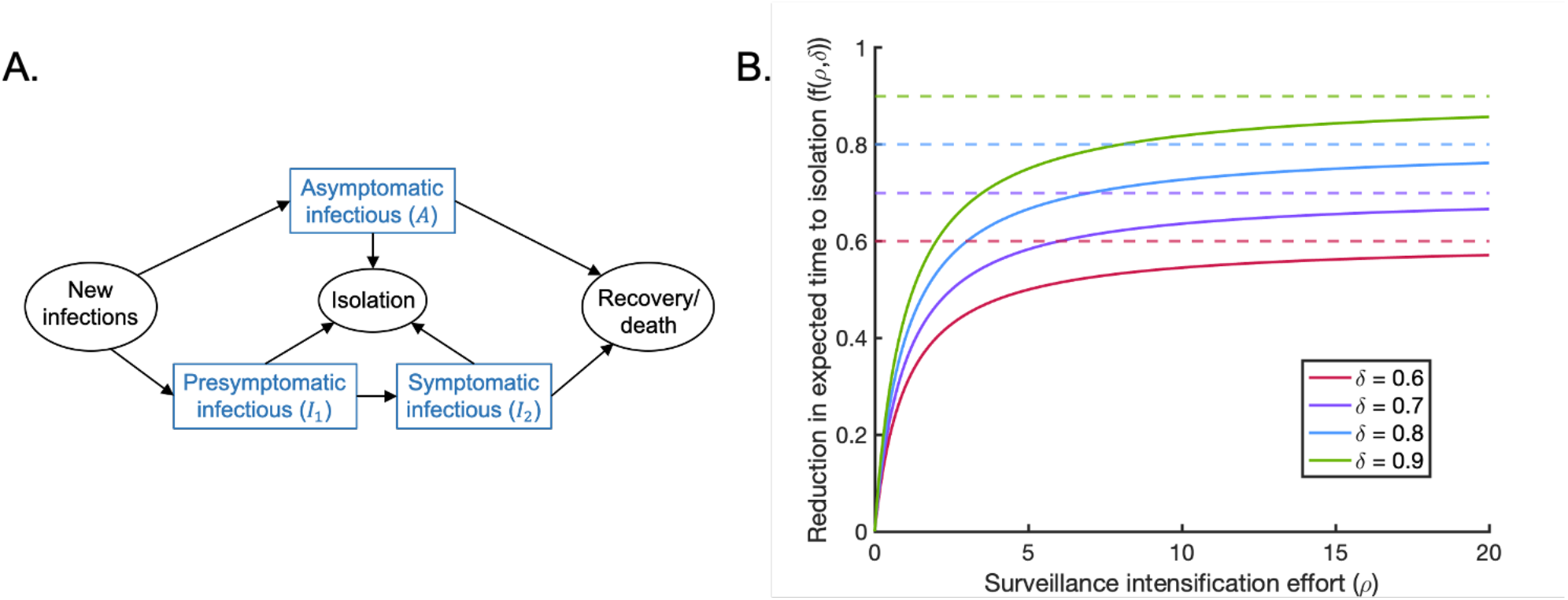
The branching process model used in our analyses. A. Schematic showing the different event types in the branching process model. The parameters of the model are described in the text and in Table 1.B. The relationship between the surveillance intensification effort (*ρ*) and the proportional reduction in the expected time to isolation (*f* (*ρ, δ*)), shown for different values of the parameter *δ* (solid lines). The parameter *δ* ∈ (0,1) represents the upper bound of *f* (*ρ, δ*) (dotted lines). This general functional relationship between surveillance effort and isolation effectiveness is assumed to hold for surveillance of both nonsymptomatic and symptomatic individuals, although nonsymptomatic hosts are more challenging to detect than symptomatic hosts (*ε* < 1).

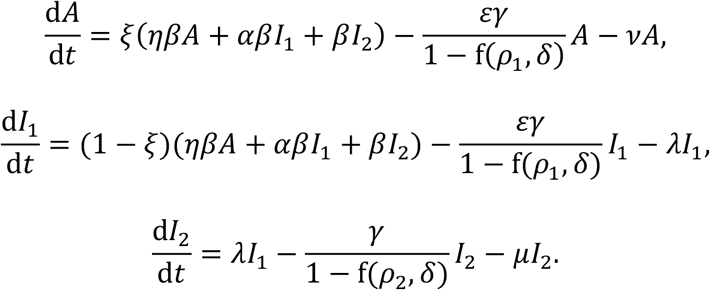

**Table 1.**
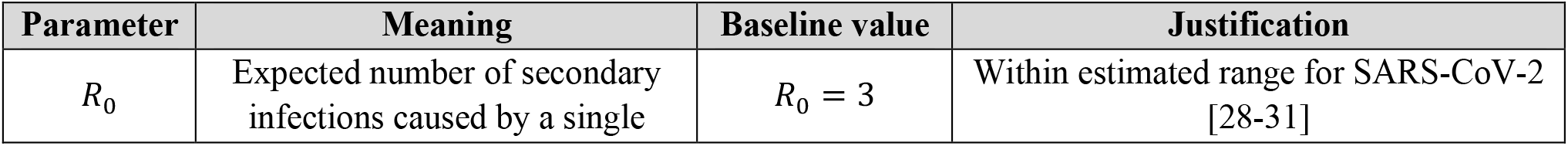
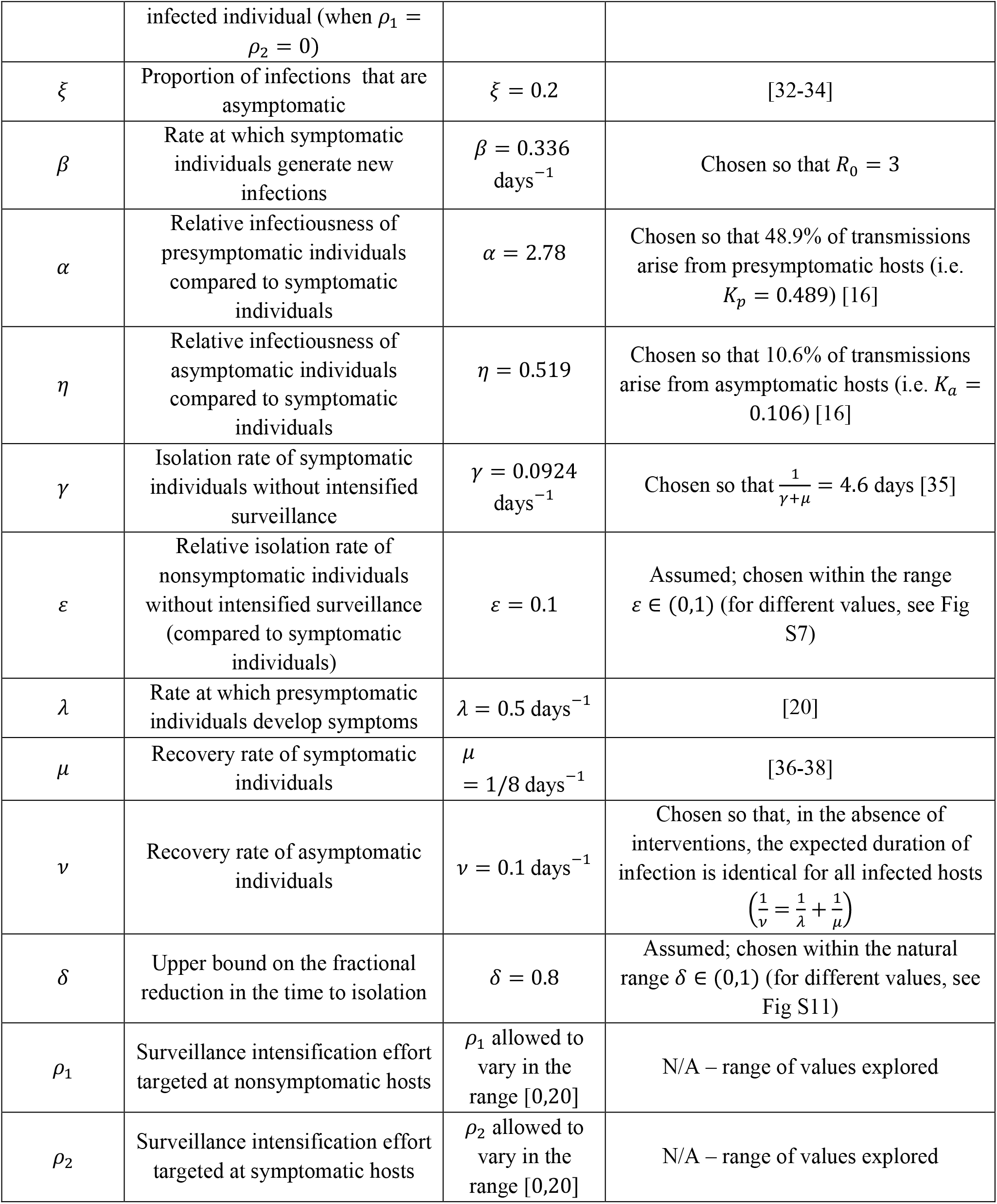
Parameters of the model and the values used in the baseline version of our analysis.

The parameters of the model, and the form of the function *f* (*ρ, δ*) that describes how the expected time to isolation is reduced for a given surveillance effort, are outlined below.

In our model, the parameter *β* and its scaled counterparts *αβ* and *ηβ* represent the rates at which symptomatic, presymptomatic and asymptomatic hosts generate new infections, respectively. Since we are modelling the beginning of a potential local outbreak, we assume that the size of the susceptible population remains approximately constant and do not track the depletion of this population. The parameter *λ* governs the rate at which presymptomatic individuals develop symptoms, so that the expected duration of the presymptomatic period is 1/*λ* days in the absence of interventions. Similarly, without interventions, the expected durations of the symptomatic and asymptomatic infectious periods are 1/*μ* days and 1/*v* days, respectively.

The baseline rate at which symptomatic individuals are detected and isolated is determined by the parameter *γ*. Assuming that nonsymptomatic individuals are more difficult to detect than symptomatic individuals, we take the analogous quantity for nonsymptomatic hosts to be *εγ*, where the scaling factor *ε* < 1 reflects the fact that interventions targeting nonsymptomatic hosts are likely to be less effective for the same surveillance effort. We assume that the sensitivity of surveillance is identical for presymptomatic and asymptomatic individuals, and therefore use the same isolation rate for both of these groups.

The parameters *ρ*_1_ and *ρ*_2_ represent the surveillance intensification effort targeted at nonsymptomatic and symptomatic hosts, respectively. The function 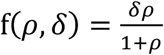 governs the proportional reduction in the expected time to isolation for a given surveillance effort, *ρ* (for a similar approach in which the proportion of infectious cases prevented is assumed to be a function of control effort, see Matthews *et al*. [27]). The functional form of *f* (*ρ, δ*) is chosen for three main reasons. First, it generates a reduced expected time to isolation when the surveillance effort increases. Second, since the proportional reduction in the expected time to isolation is bounded above by the parameter *δ* ∈ (0,1), the isolation rate saturates and cannot increase indefinitely. Third, the gradient 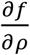 decreases with the surveillance effort *ρ*, meaning that an increase in the surveillance effort has a greater impact at low surveillance efforts compared to when this effort is already large [27]. The function *f* (*ρ, δ*) is shown in Fig 1B for different values of the parameter *δ*.

### 2.2 Reproduction number

The basic reproduction number, *R*_0_, represents the expected number of secondary infections generated by a single infected individual introduced at the start of their infection into a fully susceptible population in the absence of intensified surveillance:

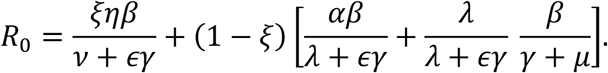

This expression is the sum of the expected number of transmissions from a host who begins in the asymptomatic class and from a host who begins in the presymptomatic infectious class, weighted by the respective probabilities *ξ* and 1 − *ξ* that determine the chance that the host experiences a fully asymptomatic course of infection. The expected number of transmissions from a host who begins in the presymptomatic infectious class comprises transmissions occurring during the incubation period and transmissions occurring during the symptomatic period, accounting for the possibility that the host is isolated prior to developing symptoms.

The proportion of infections arising from presymptomatic hosts in the absence of intensified surveillance is then given by

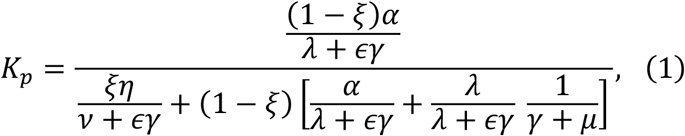

and the equivalent quantity for asymptomatic hosts is given by

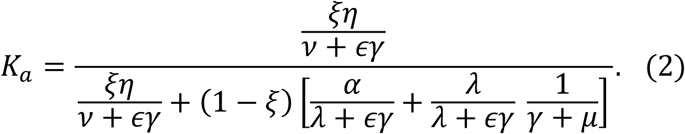

### 2.3 Baseline values of model parameters

Since this research was motivated by the need to estimate outbreak risks outside China in the initial stages of the COVID-19 pandemic, we used a baseline set of parameter values in our analyses that was informed by studies conducted during this pandemic (Table 1). Where possible, these parameter values were obtained from existing literature. However, we also performed sensitivity analyses to determine how our results varied when the parameter values were changed (see Supplementary Text S3 and Supplementary Figs S3-12). In Table 1, and throughout, rounded values are given to three significant figures.

The value of the parameter governing the baseline rate at which symptomatic individuals are isolated, *γ*, was chosen to match empirical observations which indicate that individuals who seek medical care prior to recovery or death do so around four to six days after symptom onset [35]. Specifically, we assumed that the period of time to the first medical visit could be used a proxy for the time to isolation, and chose *γ* so that the expected time period to isolation conditional on isolation occurring during the symptomatic period was given by 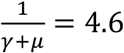 days [35]. This is different to the time period that we refer to as the expected time to isolation for symptomatic hosts, which is 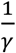 days (see Methods).

### 2.4 Probability of a local outbreak

For stochastic simulations of compartmental epidemiological models starting from a small number of hosts infected initially, there are generally two qualitatively different types of behaviour. The pathogen may fade out rapidly, or case numbers may begin to increase exponentially (only starting to fade out once the number of susceptible individuals has been sufficiently depleted, unless public health measures are introduced to reduce transmission). Consequently, running many simulations of those types of model with *R*_0_ larger than but not close to one, the epidemic size is distributed bimodally, with the total number of individuals ever infected falling into one of two distinct ranges (for a simple example, see Supplementary Fig S1A; see also [39-41]). In that scenario, a natural definition for the probability of a local outbreak is therefore the proportion of outbreak simulations for which the total number of infected individuals falls into the higher of these two ranges.

Here, since we are considering the initial phase of potential local outbreaks, we instead considered a branching process model in which depletion of susceptibles was not accounted for. If simulations of branching process models are run, then in each simulation the pathogen either fades out with few infections or case numbers generally increase indefinitely. The probability of a local outbreak starting from a small number of infected hosts then corresponds to the proportion of simulations in which the pathogen does not fade out quickly and case numbers increase indefinitely instead. This again provides a natural definition of a local outbreak, since simulations can be partitioned into two distinct sets (for an example in which simulations of a simple branching process model are used to calculate the probability of a local outbreak, see Supplementary Fig S1B).

As an alternative to repeated simulation, we instead use our branching process model (Fig 1A) to perform analytic calculations of the probability that a single imported infectious host initiates a local outbreak. To do this, we denote the probability of a local outbreak not occurring, starting from *i* presymptomatic hosts, *j* symptomatic hosts, and *k* asymptomatic hosts, by *q*_*i,j,k*_. Starting from one presymptomatic host (so that *i* = 1 and *j* = *k* = 0), there are four possibilities for the next event. That host could:

i. generate a new asymptomatic infection (with probability 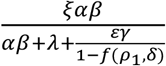);
ii. generate a new presymptomatic infection (with probability 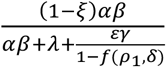);
iii. develop symptoms (with probability 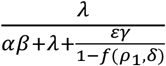), or;
iv. be isolated (with probability 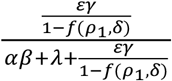).

These probabilities are obtained by considering the rates at which different possible events occur in the branching process model. Presymptomatic hosts generate new infections at rate *αβ*, and these new infections occur in asymptomatic and presymptomatic hosts with probabilities *ξ* and 1 − *ξ*, respectively. Therefore, starting from a single presymptomatic host, new asymptomatic infections occur at rate *ξαβ*, whilst new presymptomatic infections occur at rate (1 − *ξ*)*αβ*.

Additionally, presymptomatic hosts develop symptoms at rate *λ*, and are isolated at rate 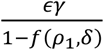.

The overall rate at which events occur is the sum of these individual event rates:

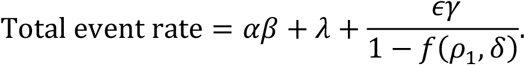

For each of the four possible next events (i-iv, above), the probability that event occurs next is the individual rate at which that event occurs divided by the total event rate, leading to the expressions given.

We use these probabilities to condition on the event that occurs next in the branching process, following the introduction of a single presymptomatic infectious individual into the population. If that event is the generation of a new asymptomatic infection, which occurs with probability 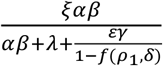, the probability that a local outbreak subsequently does not occur is *q*_1,0,1_. Applying analogous reasoning to the other possible events, we obtain

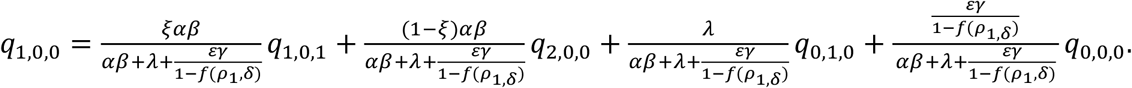

If there are no infectious hosts present in the population (i.e. *i* = *j* = *k* = 0), then a local outbreak will not occur and so *q*_0,0,0_ = 1. Assuming that transmission chains arising from two infectious individuals are independent gives *q*_1,0,1_ = *q*_1,0,0_ *q*_0,0,1_ and 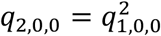. Hence,

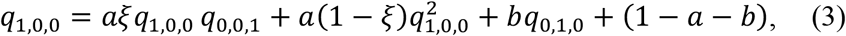

where 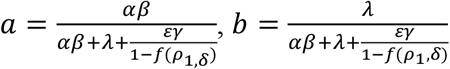.

Similarly, considering the probability of a local outbreak failing to occur starting from a single symptomatic host gives

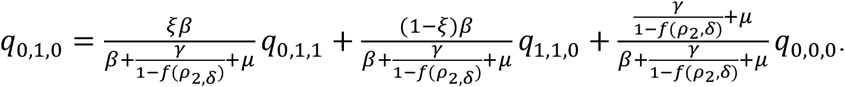

As before, noting that *q*_0,0,0_ = 1 and assuming that different infection lineages are independent leads to

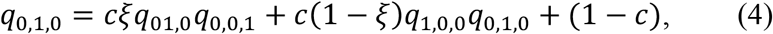

where 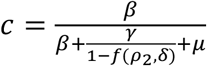.

Finally, considering the probability of a local outbreak failing to occur starting from a single asymptomatic host gives

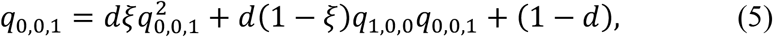

where 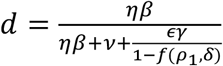.

Equations (3), (4) and (5) may be combined to give a single quartic equation for *q*_0,0,1_, yielding four sets of solutions for *q*_1,0,0_, *q*_0,1,0_ and *q*_0,0,1_ (see Supplementary Text S1). It is straightforward to verify that *q*_1,0,0_ = *q*_0,1,0_ = *q*_0,0,1_ = 1 is always a solution, and further solutions can be found numerically. The appropriate solution to take is the minimal non-negative real solution 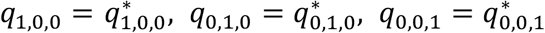 (see Supplementary Text S1). Then, the probability of a local outbreak occurring beginning from a single presymptomatic host is given by

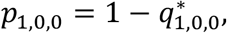

with equivalent expressions holding for *p*_0,1,0_ and *p*_0,0,1_ (the probability of a local outbreak occurring beginning from a single symptomatic host or a single asymptomatic host, respectively).

Throughout, we consider the probability *p* of a local outbreak starting from a single nonsymptomatic host entering the population, accounting for the possibility that the nonsymptomatic host is either presymptomatic or asymptomatic:

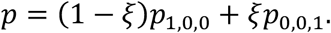

## 3. Results

### 3.1 Probability of a local outbreak

We considered the effect of *R*_0_ and the duration of the presymptomatic and asymptomatic periods on the probability of a local outbreak when a nonsymptomatic host enters a new host population (Fig 2). We examined presymptomatic periods of length 1/*λ* = 1 day, 1/*λ* = 2 days and 1/*λ* = 4 days; in each case, the duration of the asymptomatic period (1/*v* days) was adjusted so that the relative proportion of infections arising from asymptomatic hosts compared to presymptomatic hosts remained fixed (*K*_*a*_*/K*_*p*_ = 0.218, as in the baseline case). If instead nonsymptomatic infections are not accounted for, the infectious period follows an exponential distribution and the probability of a local outbreak is given by *p* = 1 − 1/*R*_0_ (red dash-dotted line in Fig 2A). Including nonsymptomatic infection in the model therefore led to an increased risk of a local outbreak in the absence of surveillance intensification (Fig 2A).

**Fig 2.**
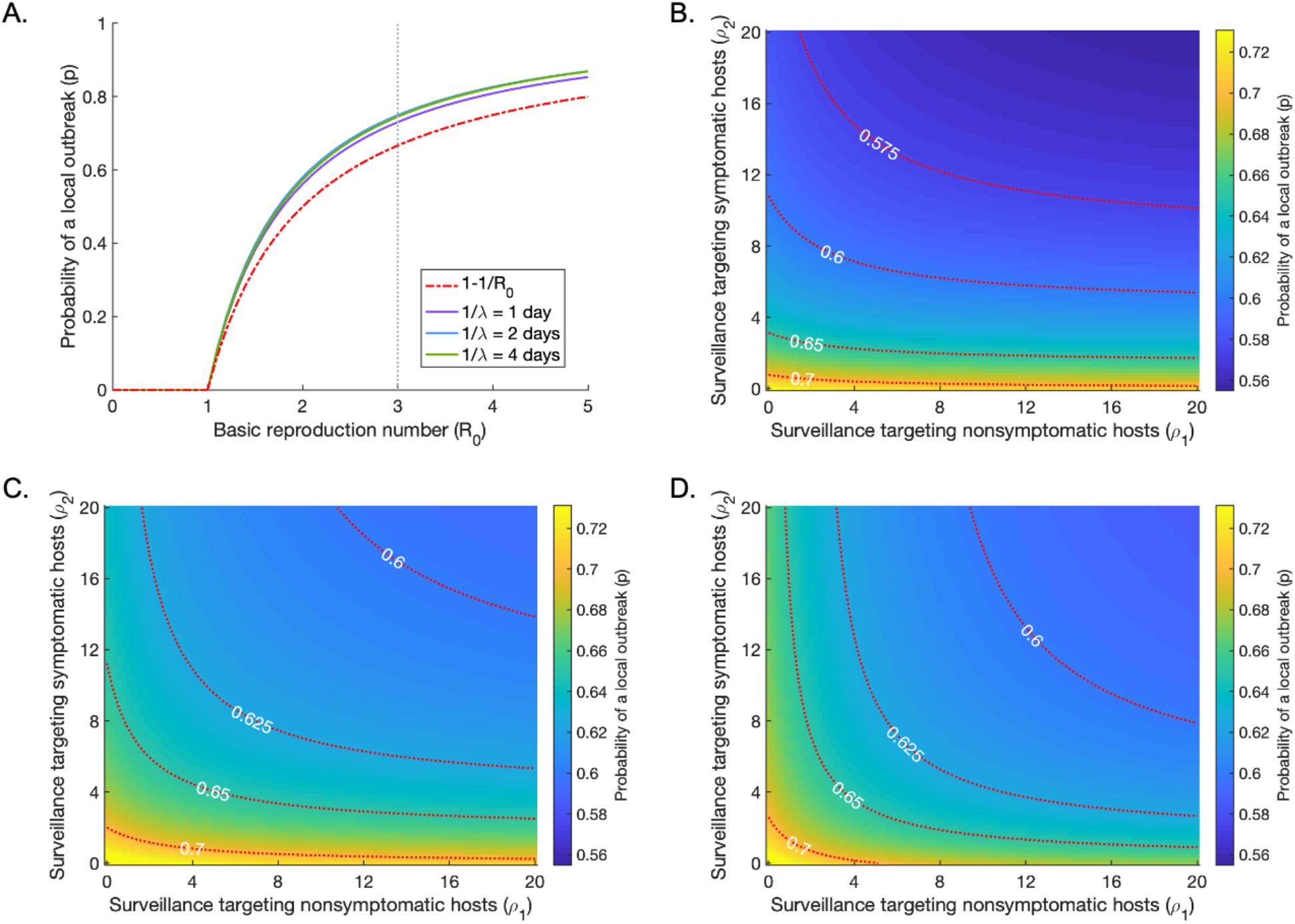
The effect of the duration of the presymptomatic and asymptomatic periods on the probability of a local outbreak (*p*), starting from a single nonsymptomatic host. A. The probability of a local outbreak as a function of the basic reproduction number *R*_0_, for presymptomatic periods of lengths 1/*λ* = 1 day (purple), 1/*λ* = 2 days (blue) and 1/*λ* = 4 days (green) in the absence of enhanced surveillance (*ρ*_1_ = *ρ*_2_ = 0). In each case, the duration of the asymptomatic period (1/*v*) is adjusted so that the relative proportion of infections arising from asymptomatic hosts compared to presymptomatic hosts remains constant (*K*_*a*_/*K*_*p*_ = 0.218, as in the baseline case). The red dash-dotted line indicates the probability of a local outbreak in the absence of nonsymptomatic transmission. The vertical grey dotted line indicates *R*_0_ = 3, the baseline value used throughout. B. The probability of a local outbreak as a function of the surveillance intensification efforts *ρ*_1_ and *ρ*_2_, for 1/*λ* = 1 day. C. The analogous figure to B but with 1/*λ* = 2 days. D. The analogous figure to B but with 1/*λ* = 4 days. Red dotted lines indicate contours of constant local outbreak probability (i.e. lines on which the probability of a local outbreak takes the values shown). The value of *β* is varied in each panel to fix *R*_0_ = 3. All other parameter values are held fixed at the values in Table 1 (except where stated).

We then considered the dependence of the probability of a local outbreak on the intensity of surveillance targeting nonsymptomatic and symptomatic hosts (Fig 2B-D). The maximum value of the surveillance intensification effort that we considered (given by *ρ*_1_ or *ρ*_2_ values of 20) corresponded to a 76% reduction in the expected time to isolation (blue line in Fig 1B), i.e. a 76% reduction in 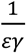 or 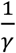.

The length of the presymptomatic and asymptomatic periods significantly affected the dependence of the probability of a local outbreak on the level of surveillance targeted at nonsymptomatic and symptomatic hosts. In Fig 2B, in which the duration of the presymptomatic period was 1 day, increasing surveillance targeted at nonsymptomatic hosts (*ρ*_1_) had a limited effect on the probability of a local outbreak, while increasing surveillance targeted at symptomatic hosts (*ρ*_2_) had a more significant effect. For example, increasing the surveillance effort targeted at nonsymptomatic hosts to *ρ*_1_ = 5 (a 67% reduction in the time to isolation) only reduced the probability of a local outbreak from 0.730 to 0.716, whereas the equivalent effort targeted at symptomatic hosts (*ρ*_2_ = 5) reduced the probability to 0.630. As shown in Figs 3C and D, however, when the presymptomatic and asymptomatic periods were longer, the benefit of directing surveillance resources towards detecting nonsymptomatic individuals increased. This was because longer presymptomatic and asymptomatic periods increased the proportion of infections generated by nonsymptomatic individuals (*K*_*p*_ + *K*_*a*_, see equations (1) and (2)); a presymptomatic period of 1 day, 2 days and 4 days corresponded to values of *K*_*p*_ + *K*_*a*_ equal to 0.424, 0.595 and 0.746, respectively.

**Fig 3.**
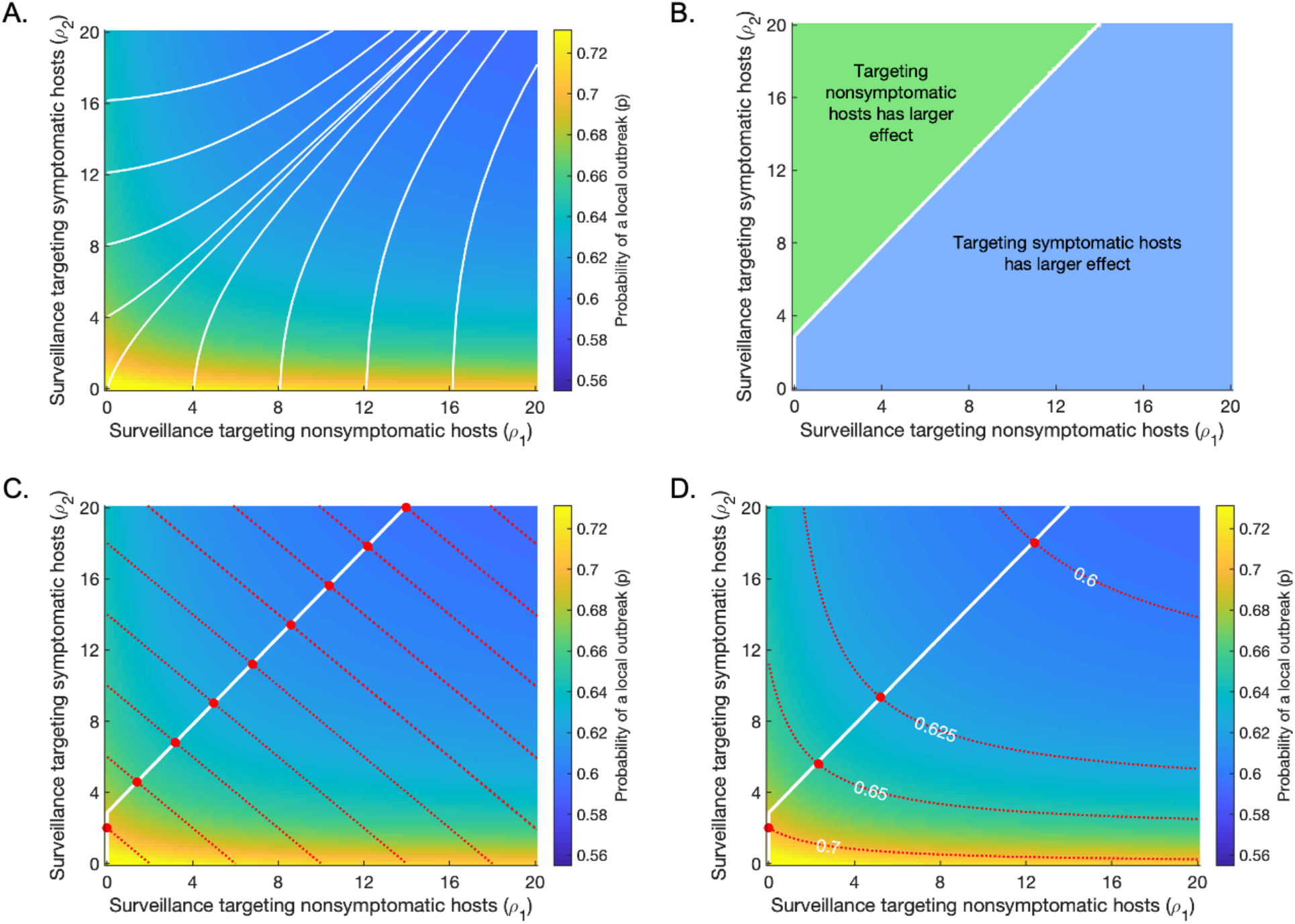
Optimal surveillance strategies to reduce the probability of a local outbreak (*p*) starting from a single nonsymptomatic host. A. The local outbreak probability for different values of *ρ*_1_ and *ρ*_2_, with the steepest descent contours overlaid (white lines). For the maximum reduction in the probability of a local outbreak at each point, surveillance must be enhanced for both nonsymptomatic and symptomatic individuals, with different levels of prioritisation depending on the current values of *ρ*_1_ and *ρ*_2_. B. Values of *ρ*_1_ and *ρ*_2_ for which increasing surveillance for nonsymptomatic hosts (i.e. increasing *ρ*_1_) is more effective at reducing the local outbreak probability than increasing surveillance for symptomatic hosts (i.e. increasing *ρ*_2_) (green region) and vice versa (blue region). The white line represents the steepest descent contour starting from *ρ*_1_ = *ρ*_2_ = 0, under the constraint that surveillance can only be enhanced for either symptomatic or nonsymptomatic hosts at any time. The diagonal section of the steepest descent contour is made up of small horizontal and vertical sections. C. Strategies for minimising the local outbreak probability for a given fixed total surveillance effort (*ρ*_1_ + *ρ*_2_ = *C*). Red dotted lines indicate contours on which *ρ*_1_ + *ρ*_2_ is constant (i.e. lines on which *ρ*_1_ + *ρ*_2_ takes the values shown); red circles indicate the points along these contours at which the local outbreak probability is minimised. The white line indicates the optimal surveillance enhancement strategy if the maximum possible surveillance level (i.e. the maximum value of *ρ*_1_ + *ρ*_2_ = *C*) is increased. D. Strategies for minimising the surveillance effort required to achieve a pre-specific risk level (an “acceptable” local outbreak probability). Red dotted lines indicate contours of constant local outbreak probability (i.e. lines on which the probability of a local outbreak takes the values shown); red circles indicate the points along these contours at which the total surveillance effort *ρ*_1_ + *ρ*_2_ is minimised. The white line indicates the optimal strategy to follow if the pre-specified risk level is increased or reduced.

### 3.2 Optimising surveillance enhancement

We next considered in more detail the impact of surveillance targeted at nonsymptomatic hosts (*ρ*_1_) relative to the impact of surveillance targeted at symptomatic hosts (*ρ*_2_). For our baseline parameter values, we considered the probability of a local outbreak starting from a single imported nonsymptomatic individual for a range of values of *ρ*_1_ and *ρ*_2_. We calculated the steepest descent contours (white lines in Fig 3A) numerically using a gradient maximisation approach, in which at each point the contour direction was determined by minimising the local outbreak probability over a fixed search radius (see Supplementary Text S2 and Supplementary Fig S2). These contours indicate how *ρ*_1_ and *ρ*_2_ should be altered to maximise the reduction in the probability of a local outbreak. In this case, enhancing surveillance targeting both symptomatic and nonsymptomatic hosts is always optimal (the steepest descent contours are neither horizontal nor vertical).

We then considered a scenario in which, at any time, it is only possible to direct resources towards enhancing surveillance of either nonsymptomatic individuals or symptomatic individuals (e.g. antigen testing of nonsymptomatic contacts of known infectious individuals, or screening for symptomatic individuals at public events). In Fig 3B, the blue region represents values of *ρ*_1_ and *ρ*_2_ for which enhancing surveillance targeting symptomatic hosts (i.e. increasing *ρ*_2_) leads to a larger reduction in the local outbreak probability than enhancing surveillance targeting nonsymptomatic hosts (i.e. increasing *ρ*_1_). In contrast, in the green region, enhancing surveillance of nonsymptomatic individuals is more effective than enhancing surveillance of symptomatic individuals. The white line represents the steepest descent contour starting from *ρ*_1_ = *ρ*_2_ = 0, under the constraint that surveillance can only be enhanced for symptomatic or nonsymptomatic hosts at any time.

Practical deployment of surveillance is often subject to logistical constraints, and policy-makers may wish to design surveillance strategies to achieve a specific objective – for example, to maximise the effectiveness of limited resources or to minimise the cost of achieving a desired outcome. We therefore also considered the following two examples of such objectives.

#### Objective 1: Minimise the probability of a local outbreak for a fixed total surveillance effort

First, we considered the question: given a fixed maximum surveillance effort (*ρ*_1_ + *ρ*_2_ = *C*), how should surveillance be targeted at nonsymptomatic and symptomatic hosts? This involves setting the values of *ρ*_1_ and *ρ*_2_ to minimise the local outbreak probability. The optimal strategies in this case are shown in Fig 3C. The red dotted lines represent contours along which the total surveillance effort *ρ*_1_ + *ρ*_2_ is held constant (i.e. different values of *C*). On each contour, the red circle indicates the point at which the local outbreak probability is minimised.

If surveillance resources are increased (i.e. *C* increases), a further question is how surveillance should then be increased. In Fig 3C, the white line represents the contour of steepest descent, under the constraint that the total change in surveillance effort (*ρ*_1_ + *ρ*_2_) is held constant at each step (rather than a constant search radius, as in Fig 3A – for more details, see Supplementary Text S2 and Supplementary Fig S2). This contour coincides exactly with that shown in Fig 3B.

These results indicate that, if surveillance resources are such that *C* is greater than 2.8 (corresponding to a 59% reduction in time to isolation of symptomatic hosts), the optimal surveillance strategy involves both enhanced surveillance of symptomatic individuals and nonsymptomatic individuals (the red dots correspond to strictly positive values of both *ρ*_1_ and *ρ*_2_, unless *C* is less than 2.8).

#### Objective 2: Minimise the total surveillance effort to achieve a pre-specified reduction in the probability of a local outbreak

Second, we considered the question: given a pre-specified acceptable risk level (i.e. probability of a local outbreak), how should the surveillance level targeted at nonsymptomatic and symptomatic hosts be chosen? This involves choosing *ρ*_1_ and *ρ*_2_ to minimise *ρ*_1_ + *ρ*_2_ along a given contour corresponding to a fixed local outbreak probability (red dotted lines in Fig 3D). On each contour, the red circle indicates the point along that contour at which the total surveillance effort *ρ*_1_ + *ρ*_2_ is minimised. These optimal points also lie exactly along the line on which enhancing surveillance targeted at symptomatic hosts is equally effective compared to enhancing surveillance targeted at nonsymptomatic hosts.

As long as the target local outbreak probability is less than 0.69, optimal surveillance involves enhanced surveillance of nonsymptomatic individuals as well as symptomatic individuals. For example, in order to reduce the local outbreak probability to 0.6, the optimal approach is to deploy resources such that *ρ*_1_ = 12.4 (a 74% reduction in time to isolation of nonsymptomatic individuals) and *ρ*_2_ = 18.0 (a 76% reduction in time to isolation of symptomatic individuals).

Plots analogous to Fig 3D in which the parameters were varied from their baseline values are shown in Supplementary Figs S3-12. In each case that we considered, our main finding was unchanged. There always exists a threshold local outbreak probability such that, if the target local outbreak probability is below this threshold, the optimal strategy for further reduction in the local outbreak probability involves enhancing surveillance targeting both nonsymptomatic and symptomatic individuals.

## 4. Discussion

A key component of infectious disease epidemic management is inferring the risk of outbreaks in different locations [5-8, 11, 41, 42]. Surveillance and control strategies can be introduced to reduce the risk that imported cases will lead to local outbreaks [12, 13, 43-46]. However, for a range of pathogens, public health measures are hindered by nonsymptomatic infectious hosts who can transmit the pathogen yet are challenging to detect [16, 42, 44, 47-49].

Here, we showed how the probability of a local outbreak can be estimated using a branching process model that accounts for transmission from nonsymptomatic infected individuals (Fig 1). The model can be used to assess the local outbreak probability for different surveillance strategies that target nonsymptomatic or symptomatic hosts (Fig 2). Previous studies have shown that detection of nonsymptomatic infections can be a key component of epidemic forecasting [42] and containment [44], and have demonstrated the benefits of identifying and isolating infectious nonsymptomatic hosts to reduce transmission [16, 17]. We focused instead on investigating how surveillance should be targeted at nonsymptomatic or symptomatic hosts in order to reduce the probability that cases imported to new locations will trigger a local outbreak (Fig 3A,B). We also showed how the optimal surveillance level targeting these two groups can be assessed when surveillance resources are limited and policy-makers have specific objectives (Fig 3C,D). In each case, our main conclusion was that surveillance for nonsymptomatic infected hosts (*ρ*_1_ > 0) can be an important component of reducing the local outbreak risk during epidemics. This result has broad implications, and our analysis could be extended to assess the potential for containing outbreaks at their source using a range of specific interventions targeting symptomatic and nonsymptomatic hosts.

Our goal here was to use the simplest possible model to explore the effects of surveillance of nonsymptomatic and symptomatic individuals on the risk of local outbreaks. However, this model is not without its limitations. One area of uncertainty is the precise values of the parameters governing pathogen transmission and control. In this article, we chose a baseline set of parameter values that is consistent with the findings of studies conducted during the COVID-19 pandemic, although constructing a detailed transmission model for this pandemic was not our main focus. For example, we set the relative rates at which presymptomatic and asymptomatic individuals generate new infections compared to symptomatic individuals so that 48.9% of transmissions arise from presymptomatic infectors, and 10.6% arise from asymptomatic infectors [16]. While this is in line with reported estimates [50, 51], there is substantial variation between studies. Similarly, the proportion of individuals who experience a fully asymptomatic course of infection (denoted by *ξ* in our model) is subject to a considerable degree of uncertainty. Here, we chose *ξ* = 0.2 as the baseline value [32-34] but estimates in the literature range from 0.04 to over 0.8 [33, 52-54]. We therefore also conducted sensitivity analyses in which we explored a range of different values of model parameters (Supplementary Text S3 and Supplementary Figs S3-12). In each case that we considered, our main conclusion was unchanged: surveillance of nonsymptomatic individuals can contribute to reducing the risk of local outbreaks. This result is expected to hold for epidemics of any pathogen for which nonsymptomatic individuals contribute significantly to transmission.

For our modelling approach to be used to make precise quantitative predictions during epidemics, it would be necessary to update the model to include the range of different specific surveillance and control interventions that are in place. For example, detection of nonsymptomatic infected individuals is facilitated by contact tracing and antigen testing, which are carried out routinely during epidemics and can be included in models explicitly [12, 44, 55, 56]. Reductions in contacts due to social distancing strategies and school or workplace closures could also be accounted for [57, 58], although such interventions are often introduced after a local outbreak has begun rather than in the initial phase of a potential local outbreak as considered here. We modelled the level of surveillance targeted at nonsymptomatic and symptomatic hosts in a simple way, using a function describing the relationship between surveillance effort and effectiveness (Fig 1B). We assumed that this general functional relationship could be applied to interventions targeting both symptomatic and nonsymptomatic hosts, accounting for logistical differences in the ease of targeting either group by scaling the effectiveness of surveillance for nonsymptomatic hosts using the parameter *ε* (results are shown for different values of *ε* in Supplementary Fig S8). In principle, it would be possible to include entirely different functional forms describing the relationship between surveillance effort and effectiveness for strategies targeting symptomatic and nonsymptomatic individuals, and these could be tailored to the effects of particular interventions. If different public health measures are included in the model explicitly, then it would be possible to increase the accuracy of assessments of the relative public health benefits of specific interventions that only target symptomatic individuals (e.g. screening for passengers with heightened temperatures at airports [59, 60]) compared to interventions that also target nonsymptomatic hosts (e.g. travel bans or quarantine of all inbound passengers [61, 62]). Of course, this would require data from which the relative effectiveness of different measures could be inferred.

The underlying transmission model could also be extended to include additional realism in several ways. Transmission dynamics are influenced by marked heterogeneities in the patterns of contacts between individuals in different age groups [63, 64], and, for COVID-19, susceptibility to infection, the likelihood of developing symptoms, and the average severity of those symptoms increase with age [65, 66]. Age-dependent variation in the proportion of asymptomatic cases in particular implies that the optimal balance of surveillance between symptomatic and nonsymptomatic hosts may differ between age groups. An age-structured version of the model presented here is a focus of our ongoing research. Similarly, for a range of infectious diseases, the distribution characterising the number of secondary infections generated by each infected host (the offspring distribution) exhibits a high degree of overdispersion [67-70]. For a fixed value of *R*_0_, a higher degree of overdispersion increases the likelihood that initial cases will fade out without leading to a local outbreak [71, 72], and suggests that greater reductions in local outbreak risks could theoretically be achieved for the same surveillance effort, if potential superspreaders or superspreading events can be identified and targeted.

Despite the necessary simplifications, we have shown how the risk of local outbreaks can be estimated during epidemics using a branching process model that includes nonsymptomatic infectious hosts explicitly. Determining the extent to which nonsymptomatic individuals contribute to transmission is essential early in emerging epidemics of a novel pathogen. As we have shown, if transmissions occur from nonsymptomatic infectors, dedicating surveillance resources towards finding nonsymptomatic cases can be an important component of public health measures that aim to prevent local outbreaks.

## Data Availability

This study did not include the collection of any data. All data referenced in the manuscript are accessible through the relevant citations.

## Acknowledgements

Thank you to Andrew Wood and Charles Tolkien-Gillett for proofreading.

## Supplementary Text

### Text S1. Probability of a local outbreak

In the Methods section of the main text, we outlined an approach for deriving the probability of a local outbreak starting from a single infectious host in either the presymptomatic, symptomatic or asymptomatic classes. Here, we provide more details about that derivation. The probability of a local outbreak not occurring, starting from *i* presymptomatic hosts, *j* symptomatic hosts and *k* asymptomatic hosts, is denoted by *q*_*i,j,k*_. If we consider the temporal evolution of (*i, j, k*) to be a Markov process on the state space *M* = *ℤ* _≥0_ × *ℤ* _≥0_ × *ℤ* _≥0_, then *q*_*i,j,k*_ is the hitting probability of the state (0,0,0) starting from the state (*i, j, k*). The vector of hitting probabilities 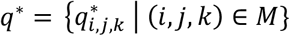 is therefore the minimal non-negative (real) solution to the following system of equations: *q*_0,0,0_ = 1,

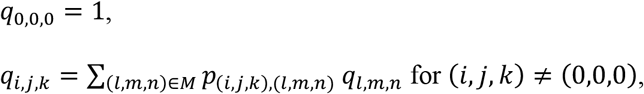

where *p*_(*i,j,k*),(*l,m,n*)_ is the transition probability from state (*i, j, k*) to state (*l, m, n*) [73].

Here, minimality means that if 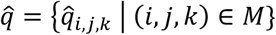 is another non-negative real solution, then 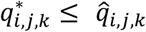 for all (*i, j, k*) ∈ *M*.

From this, equations (3), (4) and (5) in the main text are obtained. These equations may be reduced to the following quartic equation for *q*_0,0,1_:

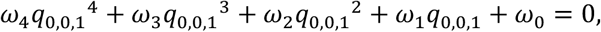

where

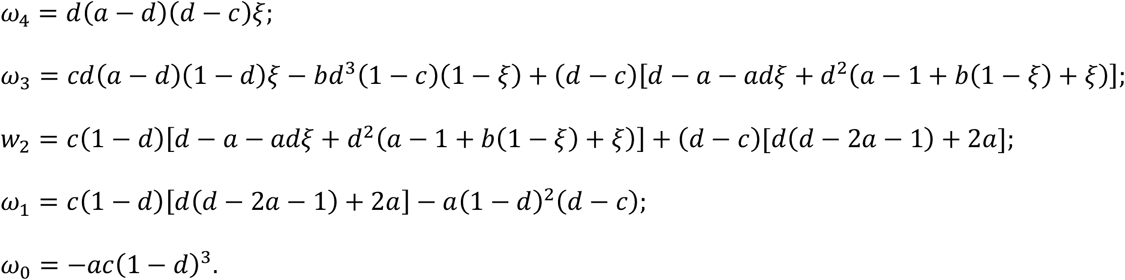

The parameters *a, b, c, d* and *ξ* are as defined in the main text.

This yields four solutions for *q*_0,0,1_ and four corresponding solutions for each of *q*_1,0,0_ and *q*_0,1,0_. One solution is always given by *q*_1,0,0_ = *q*_0,1,0_ = *q*_0,0,1_ = 1; the other solutions may be found numerically. As described above, we take the minimal non-negative real solution 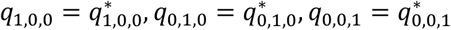, and observe that the probability of a local outbreak occurring starting from *i* presymptomatic hosts, *j* symptomatic hosts and *k* asymptomatic hosts is simply 1 – *q*_*i,j,k*_, giving the result stated in the main text.

If required, this result can be confirmed for specific model parameter values via repeated stochastic simulation of the branching process model, starting from *i* presymptomatic hosts, *j* symptomatic hosts and *k* asymptomatic hosts. As described in the main text, in simulations of branching process models, initial cases typically either fade out or go on to cause a local outbreak. There is a natural definition of a local outbreak as a simulation in which a large number of infections occur (see Supplementary Fig S1B). The probability of a local outbreak then corresponds to the proportion of simulations in which large numbers of infections occur.

We note here that although this is a natural way to define a local outbreak, alternative definitions exist that may be more appropriate in particular contexts. This is discussed by Thompson *et al*. (reference [41] in the main text), who consider three practically relevant definitions of an outbreak based on different criteria for measuring severity.

### Text S2. Computation of steepest descent contours

The steepest descent contours shown in Fig 3A of the main text were computed using a gradient maximisation approach, in which at each point the contour direction was determined by minimising the local outbreak probability over a fixed search radius (Fig S2 A,B). Starting from *ρ*_1_, *ρ*_2_, at each step we considered increasing *ρ*_1_ by an amount Δ*ρ*_1_ and increasing *ρ*_2_ by an amount Δ*ρ*_2_ subject to the constraint (Δ*ρ*_1_)^2^ + (Δ*ρ*_2_)^2^ = *r*^2^, where *r* is a small pre-specified constant. To achieve this, we scanned over the circular arc Δ*ρ*_1_ = *r* cos *θ*, Δ*ρ*_2_ = *r* s*i*n *θ*, for *θ* ∈ [0, *π*/2] (Fig S2 A). In practice, this range was discretised into 33 search directions evenly spaced between 0 and *π*/2. We then selected the pair of Δ*ρ*_1_, Δ*ρ*_2_ values for which the local outbreak probability evaluated at *ρ*_1_ + Δ*ρ*_1_, *ρ*_2_ + Δ*ρ*_2_ was minimised. The process was then repeated beginning from 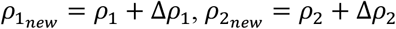 (Fig S2 B). The white line in Fig 3B, which divides the region in which increasing *ρ*_1_ has a greater effect on the local outbreak probability from the region in which increasing *ρ*_2_ has a greater effect on the local outbreak probability, was computed in an analogous way, with the additional restriction that we only considered the search directions *θ* = 0 and *θ* = *π*/2 (i.e. intensifying only surveillance of nonsymptomatic or symptomatic hosts; see Fig S2 C).

In Fig 3C, the white line represents the contour of steepest descent under the constraint that the total change in surveillance effort (Δ*ρ*_1_ + Δ*ρ*_2_ = *S*) is held constant at each step, rather than fixing the search radius (Δ*ρ*_1_)^2^ + (Δ*ρ*_2_)^2^ = *r*^2^ as in Fig 3A. Therefore, instead of scanning over a circular arc, at each step we scan along the line Δ*ρ*_1_ = *s*, Δ*ρ*_2_ = *S* − *s*, where *c* varies in the range [0, *S*] (Fig S2 D). Otherwise, the process is completely analogous to that described above.

### Text S3. Robustness of results to parameter values used

We conducted supplementary analyses to investigate how our results are affected by varying the parameters from their baseline values given in Table 1. We performed sensitivity analyses on the values of *R*_0_, *ξ, K*_*p*_, *K*_*a*_, *γ* + *μ, ϵ, λ, v* and *δ*. For each of these, we present plots analogous to Fig 3D for six different values of the relevant parameter (Figs S3-S12). In each case we considered, our qualitative message was unchanged: whenever the maximum acceptable risk level was below a particular threshold value, the optimal strategy involved surveillance targeting both nonsymptomatic and symptomatic individuals.

### Text S4. Details on computer code

All computer code was written in the MATLAB programming environment (version R2019a).

## Supplementary Figures

**Fig S1.**
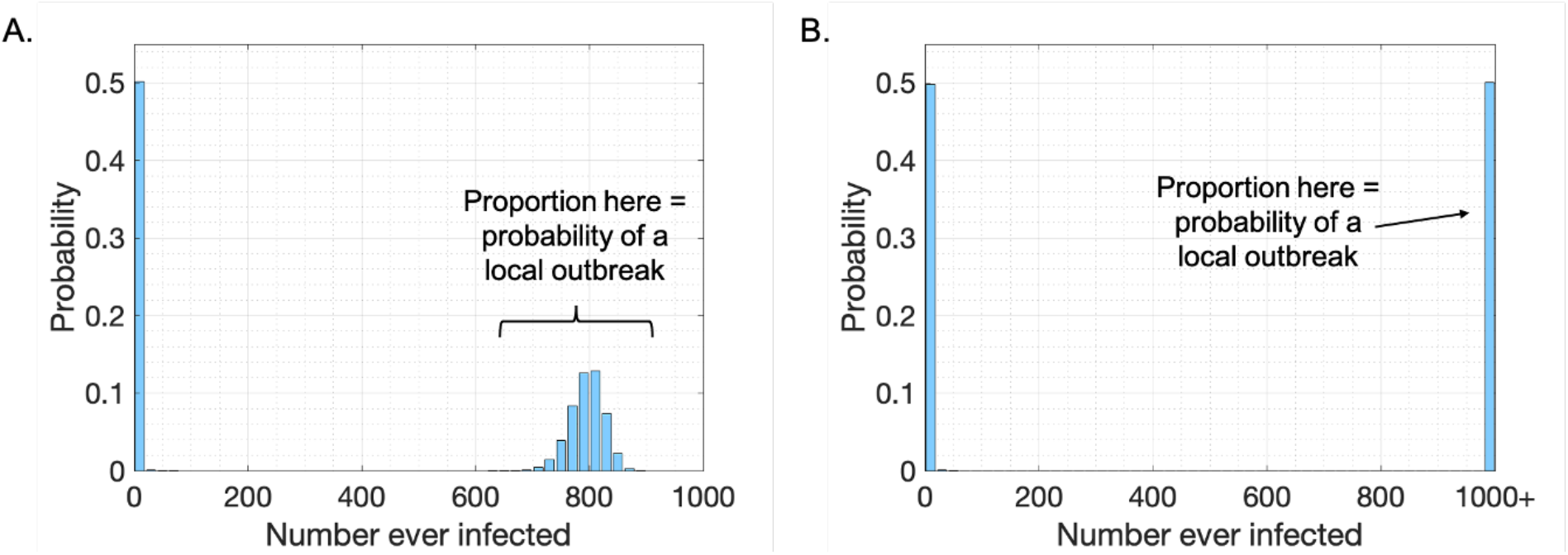
Illustrating the definition of the term ‘local outbreak’ with a simple example. A. The total number of individuals ever infected (final epidemic size) in each of 100,000 simulations of a stochastic SIR (Susceptible-Infected-Removed) model with basic reproduction number *R*_0_ = 2, beginning from a single infectious host each time. Within each simulation, each event is either an infection event (with probability 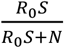) or a removal event (with probability 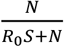), and simulations are run until the pathogen fades out (*I* hits zero). This provides a natural partitioning between simulations that fade out quickly and those that go on to become local outbreaks. In 50% of simulations, fewer than 20 infections occurred in total (left hand peak); initial cases did not lead to sustained transmission in the population. In the remaining 50% of simulations (local outbreaks), between 620 and 900 individuals were infected in total each time. The probability of a local outbreak is then defined as the proportion of simulations for which the final epidemic size lies within this natural upper range, here equal to 0.5. B. The analogous figure to panel A, but for the branching process version of the SIR model in which depletion of susceptibles is not accounted for (i.e. *S* = *N* throughout each simulation). In this model, each event is either an infection event (with probability 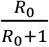) or a removal event (with probability 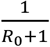), and simulations are run until either the pathogen fades out (*I* hits zero) or 1000 infections have occurred. The rightmost bar corresponds to simulations in which 1000 infections occurred. There is again a natural partitioning between simulations that fade out quickly and those that go on to become local outbreaks, with the probability of a local outbreak matching the equivalent value for the stochastic SIR model (panel A).

**Fig S2.**
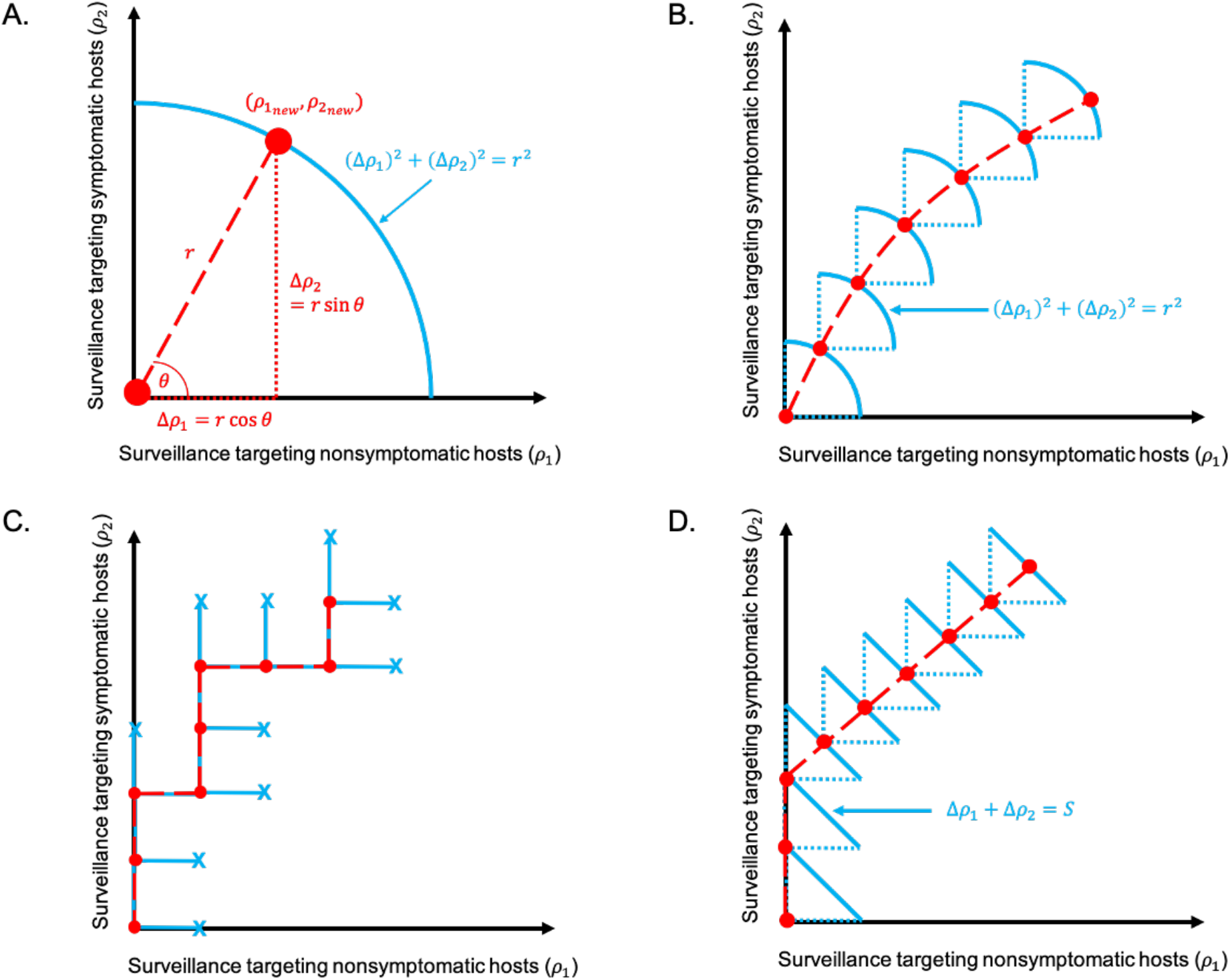
Computation of the steepest descent contours shown in the main text. A. To compute the steepest descent contours shown in Fig 3A of the main text, we increment *ρ*_1_ and *ρ*_2_ by scanning over a constant search radius (Δ*ρ*_1_)^2^ + (Δ*ρ*_2_)^2^ = *r*^2^ (blue arc), and moving to the point 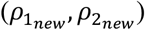 along that arc at which the local outbreak probability is minimised. B. The process shown in A is repeated to generate the complete contour (red dashed line). C. The analogous figure to B, in which the search direction is limited to directly to the right (*θ* = 0) or directly upwards (*θ* = *π*/2). This procedure is used to generate the contour in Fig 3B in the main text. D. The analogous figure to B, in which the total change in surveillance effort (Δ*ρ*_1_ + Δ*ρ*_2_ = *S*) is held constant at each step, rather than the search radius. This procedure is used to generate the contour in Figs 3C and D in the main text.

**Fig S3.**
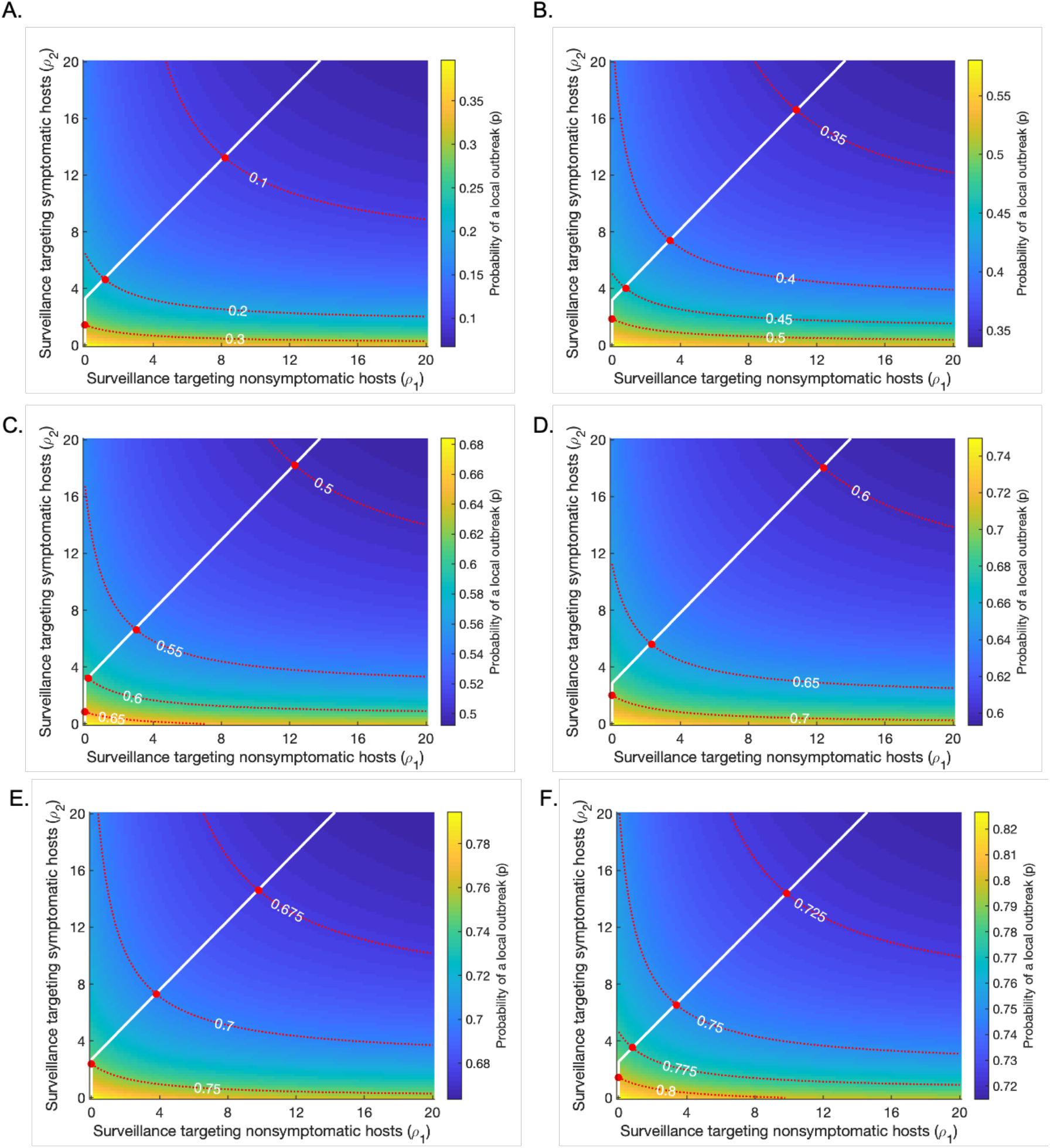
Varying the basic reproduction number *R*_0_ from its baseline value (*R*_0_ = 3). Plots are analogous to Fig 3D in the main text, showing strategies for minimising the surveillance effort required to achieve a pre-specified risk level (an “acceptable” local outbreak probability). Red dotted lines represent contours along which the probability of a local outbreak is constant, as labelled; red circles indicate the points along these contours at which the total surveillance effort *ρ*_1_ + *ρ*_2_ is minimised. The white line indicates the optimal strategy to follow if the pre-specified risk level is reduced. Apart from *R*_0_ and *β* (which is changed in each panel to set the value of *R*_0_), all parameters are held fixed at their baseline values given in Table 1. A. *R*_0_ = 1.5. B. *R*_0_ = 2. C. *R*_0_ = 2.5.D. *R*_0_ = 3 (baseline). E. *R*_0_ = 3.5. F. *R*_0_ = 4.

**Fig S4.**
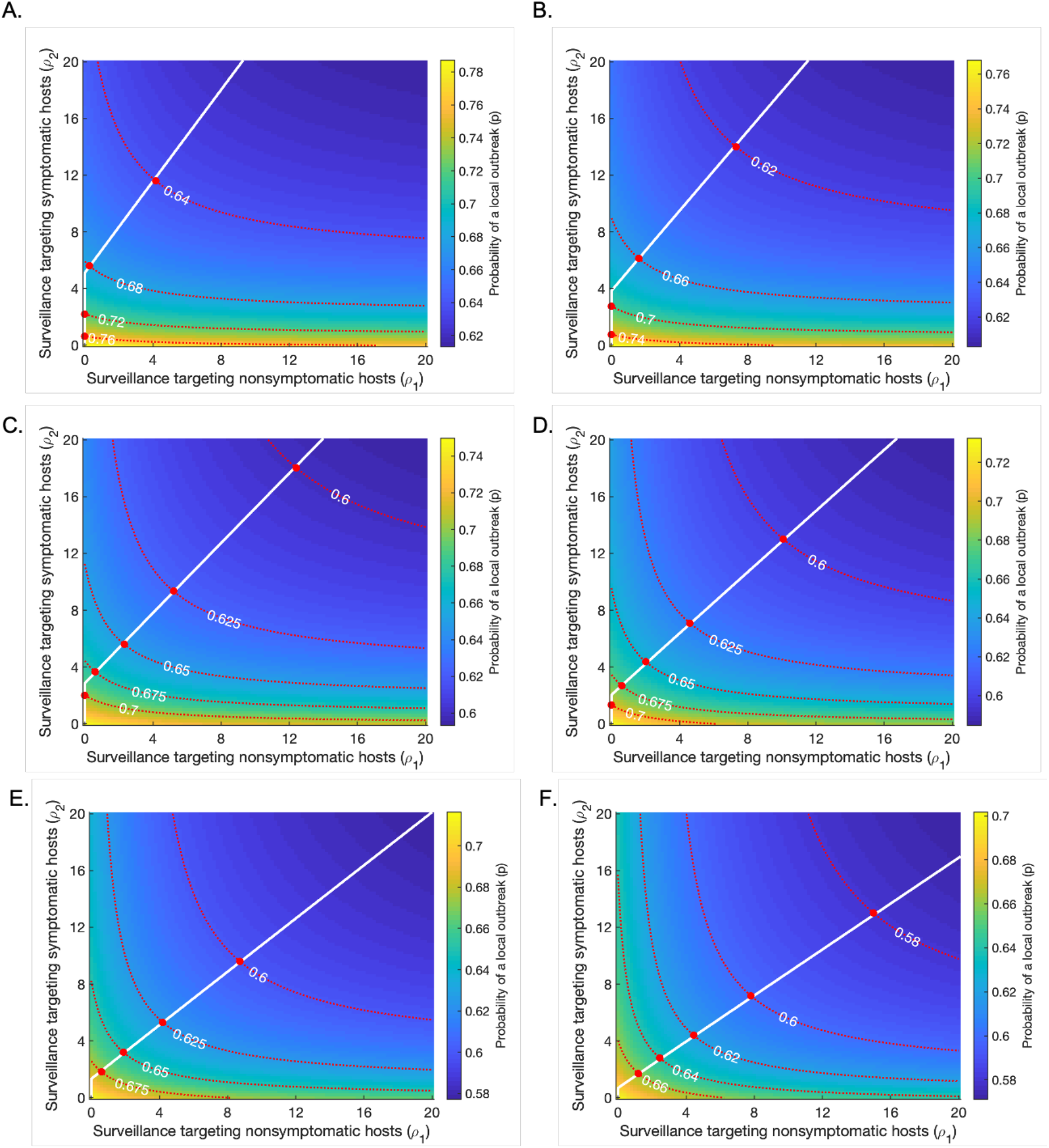
Varying the proportion of infections from asymptomatic infectors, *ξ*, from its baseline value (*ξ* = 0. 2). Plots are analogous to Fig 3D in the main text, showing strategies for minimising the surveillance effort required to achieve a pre-specified risk level (an “acceptable” local outbreak probability). Red dotted lines represent contours along which the probability of a local outbreak is constant, as labelled; red circles indicate the points along these contours at which the total surveillance effort *ρ*_1_ + *ρ*_2_ is minimised. The white line indicates the optimal strategy to follow if the pre-specified risk level is reduced. Apart from *ξ* and *β* (which is changed in each panel to set *R*_0_ = 3), all parameters are held fixed at their baseline values given in Table 1. A. *ξ* = 0. B. *ξ* = 0.1. C. *ξ* = 0.2 (baseline). D. *ξ* = 0.3. E. *ξ* = 0.4. F. *ξ* = 0.5.

**Fig S5.**
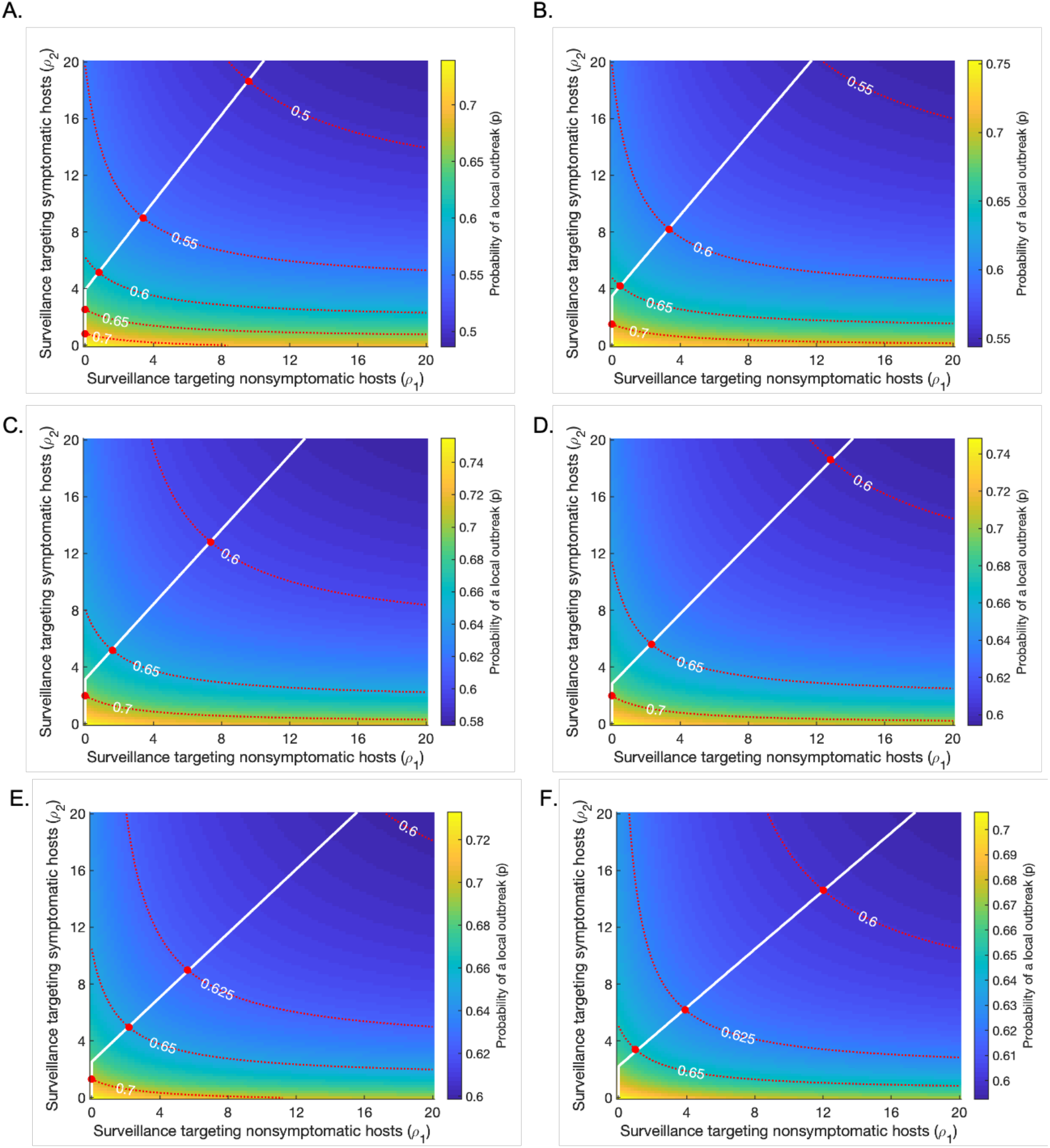
Varying the proportion of infections arising from presymptomatic hosts in the absence of intensified surveillance (*K*_*p*_, given by expression (1) in the main text) from its baseline value (*K*_*p*_ = 0. 489). In each case, the proportions of infections arising from asymptomatic and symptomatic hosts are adjusted so that they remain in the same ratio as in the baseline case. Plots are analogous to Fig 3D in the main text, showing strategies for minimising the surveillance effort required to achieve a pre-specified risk level (an “acceptable” local outbreak probability). Red dotted lines represent contours along which the probability of a local outbreak is constant, as labelled; red circles indicate the points along these contours at which the total surveillance effort *ρ*_1_ + *ρ*_2_ is minimised. The white line indicates the optimal strategy to follow if the pre-specified risk level is reduced. Apart from *K*_*p*_ and *K*_*a*_, as well as *α* and *η* (which are changed in each panel to set the values of *K*_*p*_ and *K*_*a*_), and *β* (which is changed in each panel to set *R*_0_ = 3), all parameters are held fixed at their baseline values given in Table 1. A. *K*_*p*_ = 0.2. B. *K*_*p*_ = 0.3. C. *K*_*p*_ = 0.4. D. *K*_*p*_ = 0.5. E. *K*_*p*_ = 0.6. F. *K*_*p*_ = 0.7.

**Fig S6.**
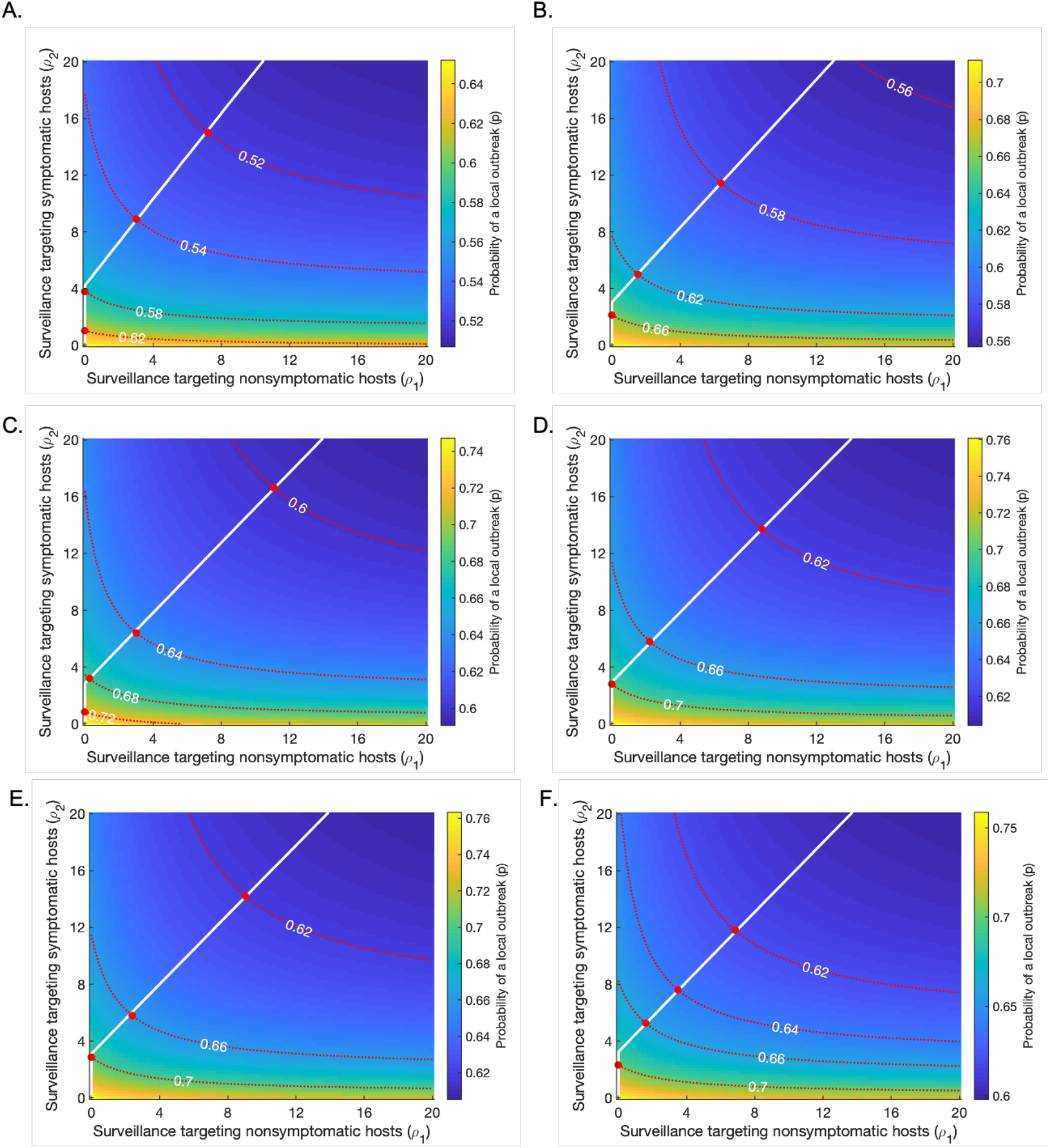
Varying the proportion of infections arising from asymptomatic hosts in the absence of intensified surveillance (*K*_a_, given by expression (2) in the main text) from its baseline value (*K*_a_ = 0. 106). In each case, the proportions of infections arising from presymptomatic and symptomatic hosts are adjusted so that they remain in the same ratio as in the baseline case. Plots are analogous to Fig 3D in the main text, showing strategies for minimising the surveillance effort required to achieve a pre-specified risk level (an “acceptable” local outbreak probability). Red dotted lines represent contours along which the probability of a local outbreak is constant, as labelled; red circles indicate the points along these contours at which the total surveillance effort *ρ*_1_ + *ρ*_2_ is minimised. The white line indicates the optimal strategy to follow if the pre-specified risk level is reduced. Apart from *K*_*p*_ and *K*_*a*_, as well as *α* and *η* (which are changed in each panel to set the values of *K*_*p*_ and *K*_*a*_), and *β* (which is changed in each panel to set *R*_0_ = 3), all parameters are held fixed at their baseline values given in Table 1. A. *K*_*a*_ = 0.01. B. *K*_*a*_ = 0.05. C. *K*_*a*_ = 0.1. D. *K*_*a*_ = 0.15. E. *K*_*a*_ = 0.2. F. *K*_*a*_ = 0.25.

**Fig S7.**
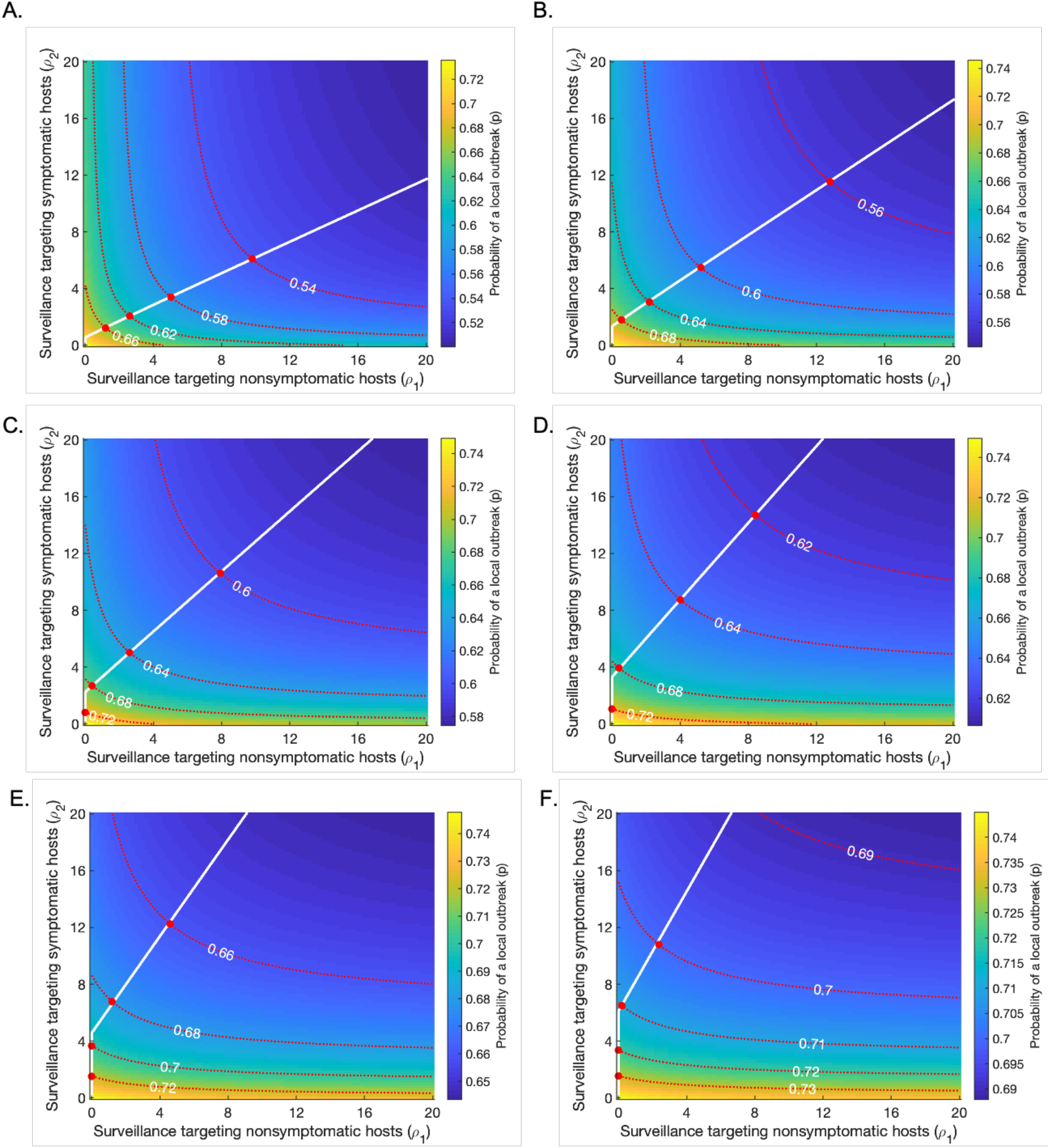
Varying the expected time period to isolation conditional on isolation occurring during the symptomatic period, 1/(*γ* + *μ*), from its baseline value (1/(*γ* + *μ*) = 4. 6 days). This is achieved by varying the parameter *γ*, whilst holding the recovery rate of symptomatic individuals *μ* equal to its baseline value (*μ* = 1/8 days ^−1^). Plots are analogous to Fig 3D in the main text, showing strategies for minimising the surveillance effort required to achieve a pre-specified risk level (an “acceptable” local outbreak probability). Red dotted lines represent contours along which the probability of a local outbreak is constant, as labelled; red circles indicate the points along these contours at which the total surveillance effort *ρ*_1_ + *ρ*_2_ is minimised. The white line indicates the optimal strategy to follow if the pre-specified risk level is reduced. Apart from *γ* and *β* (which is changed in each panel to set *R*_0_ = 3), all parameters are held fixed at their baseline values given in Table 1. A. 1/(*γ* + *μ*) = 2 days. B. 1/(*γ* + *μ*) = 3 days. C. 1/(*γ* + *μ*) = 4 days. D. 1/(*γ* + *μ*) = 5 days. E. 1/(*γ* + *μ*) = 6 days. F. 1/(*γ* + *μ*) = 7 days.

**Fig S8.**
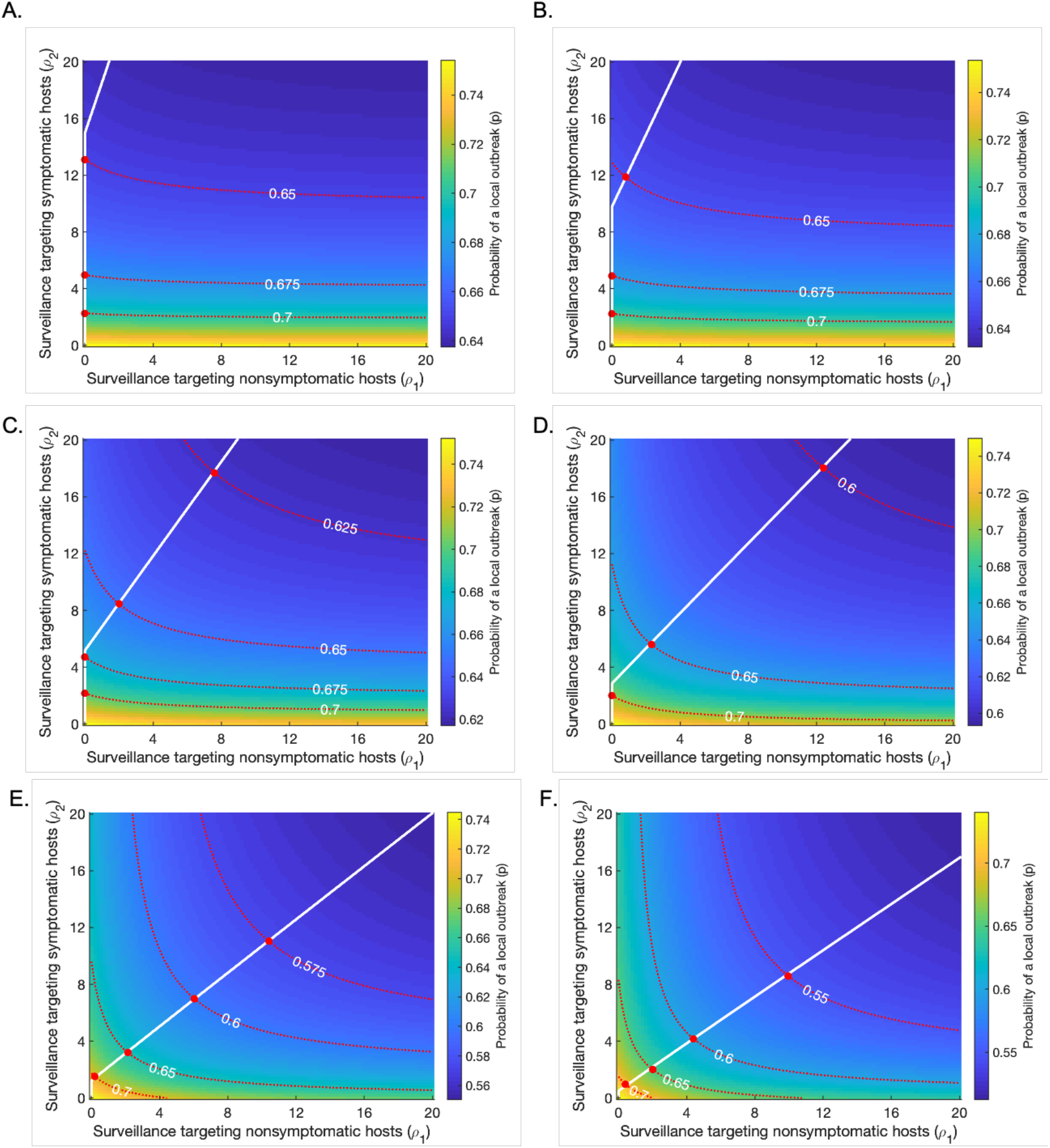
Varying *ϵ*, the relative isolation rate of nonsymptomatic individuals without intensified surveillance (compared to symptomatic individuals), from its baseline value (*ϵ* = 0. 1). Plots are analogous to Fig 3D in the main text, showing strategies for minimising the surveillance effort required to achieve a pre-specified risk level (an “acceptable” local outbreak probability). Red dotted lines represent contours along which the probability of a local outbreak is constant, as labelled; red circles indicate the points along these contours at which the total surveillance effort *ρ*_1_ + *ρ*_2_ is minimised. The white line indicates the optimal strategy to follow if the pre-specified risk level is reduced. Apart from *ϵ* and *β* (which is changed in each panel to set *R*_0_ = 3), all parameters are held fixed at their baseline values given in Table 1. A. *ϵ* = 0.01. B. *ϵ* = 0.02. C. *ϵ* = 0.05. D. *ϵ* = 0.1 (baseline). E. *ϵ* = 0.2. F. *ϵ* = 0.3.

**Fig S9.**
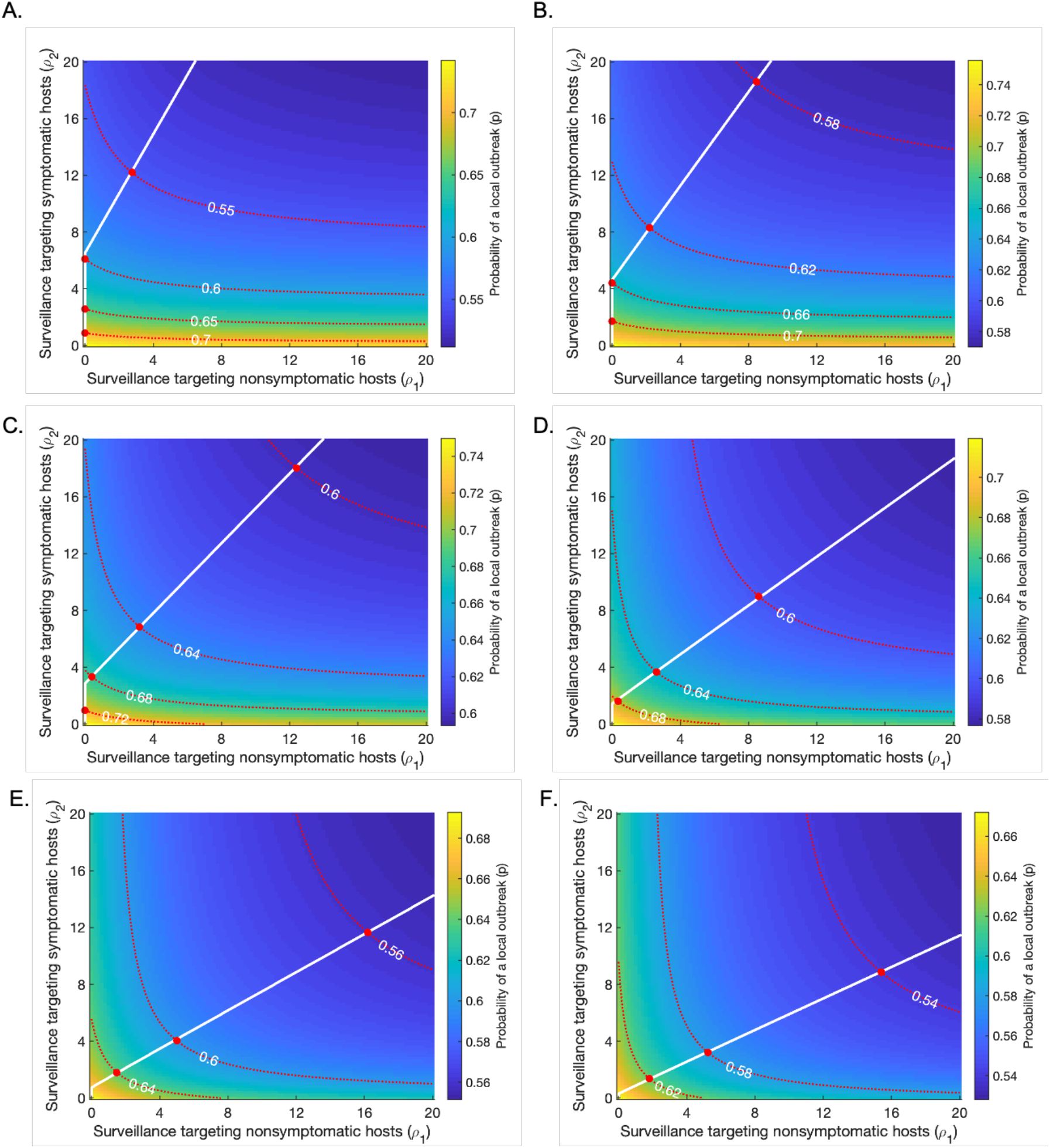
Varying the duration of the presymptomatic period, 1/*λ*, from its baseline value (1/*λ* = 0. 5 days). Plots are analogous to Fig 3D in the main text, showing strategies for minimising the surveillance effort required to achieve a pre-specified risk level (an “acceptable” local outbreak probability). Red dotted lines represent contours along which the probability of a local outbreak is constant, as labelled; red circles indicate the points along these contours at which the total surveillance effort *ρ*_1_ + *ρ*_2_ is minimised. The white line indicates the optimal strategy to follow if the pre-specified risk level is reduced. Apart from *λ* and *β* (which is changed in each panel to set *R*_0_ = 3), all parameters are held fixed at their baseline values given in Table 1. A. 1/*λ* = 0.5 days. B. 1/*λ* = 1 day. C. 1/*λ* = 2 days (baseline). D. 1/*λ* = 4 days. E. 1/*λ* = 6 days. F. 1/*λ* = 8 days.

**Fig S10.**
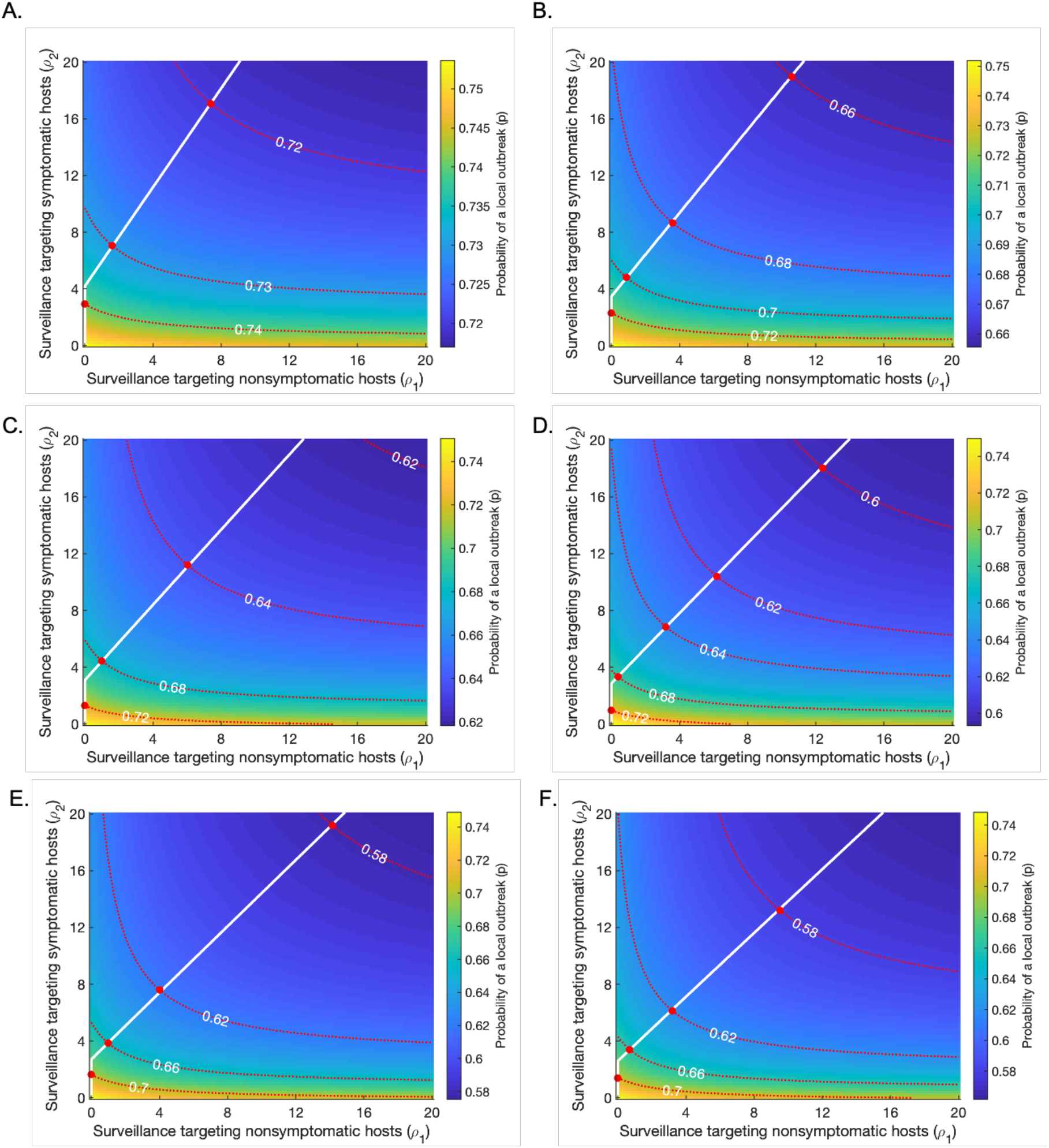
Varying the duration of the symptomatic period, 1/*μ*, from its baseline value (1/*μ* = 8 days). The parameter *γ* is varied simultaneously such that 1/(*γ* + *μ*), the expected time period to isolation conditional on isolation occurring during the symptomatic period, remains at its baseline value (1/(*γ* + *μ*) = 4.6 days). Plots are analogous to Fig 3D in the main text, showing strategies for minimising the surveillance effort required to achieve a pre-specified risk level (an “acceptable” local outbreak probability). Red dotted lines represent contours along which the probability of a local outbreak is constant, as labelled; red circles indicate the points along these contours at which the total surveillance effort *ρ*_1_ + *ρ*_2_ is minimised. The white line indicates the optimal strategy to follow if the pre-specified risk level is reduced. Apart from *μ* and *γ*, and *β* (which is changed in each panel to set *R*_0_ = 3), all parameters are held fixed at their baseline values given in Table 1. A. 1/*μ* = 5 days. B. 1/*μ* = 6 days. C. 1/*μ* = 7 days. D. 1/*μ* = 8 days (baseline). E. 1/*μ* = 9 days. F. 1/*μ* = 10 days.

**Fig S11.**
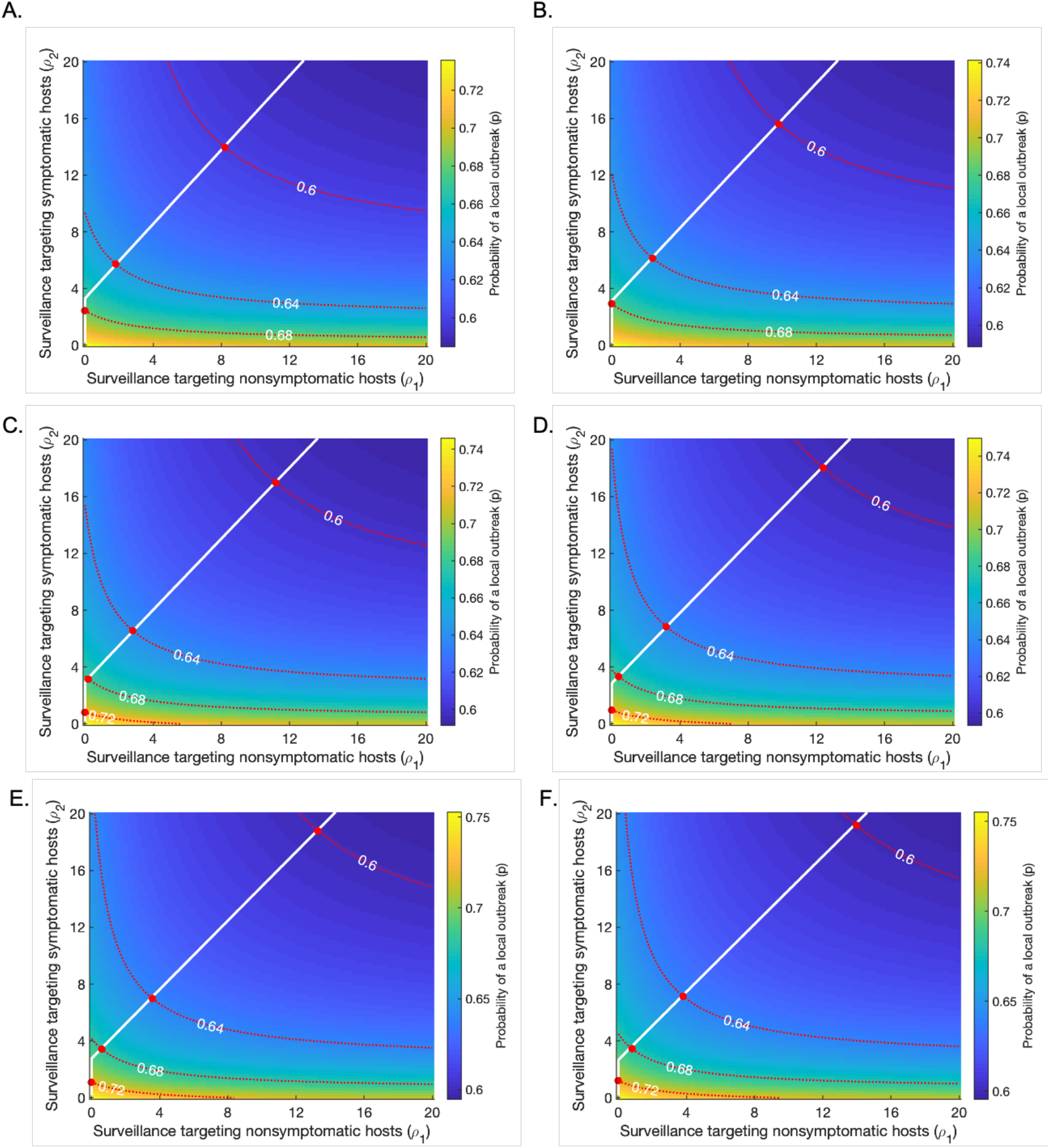
Varying 1/*v*, the duration of the asymptomatic period, from its baseline value (1/*v* = 10 days). Plots are analogous to Fig 3D in the main text, showing strategies for minimising the surveillance effort required to achieve a pre-specified risk level (an “acceptable” local outbreak probability). Red dotted lines represent contours along which the probability of a local outbreak is constant, as labelled; red circles indicate the points along these contours at which the total surveillance effort *ρ*_1_ + *ρ*_2_ is minimised. The white line indicates the optimal strategy to follow if the pre-specified risk level is reduced. Apart from *v* and *β* (which is changed in each panel to set *R*_0_ = 3), all parameters are held fixed at their baseline values given in Table 1. A. 1/*v* = 7 days. B. 1/*v* = 8 days. C. 1/*v* = 9 days. D. 1/*v* = 10 days (baseline). E. 1/*v* = 11 days. F. 1/*v* = 12 days.

**Fig S12.**
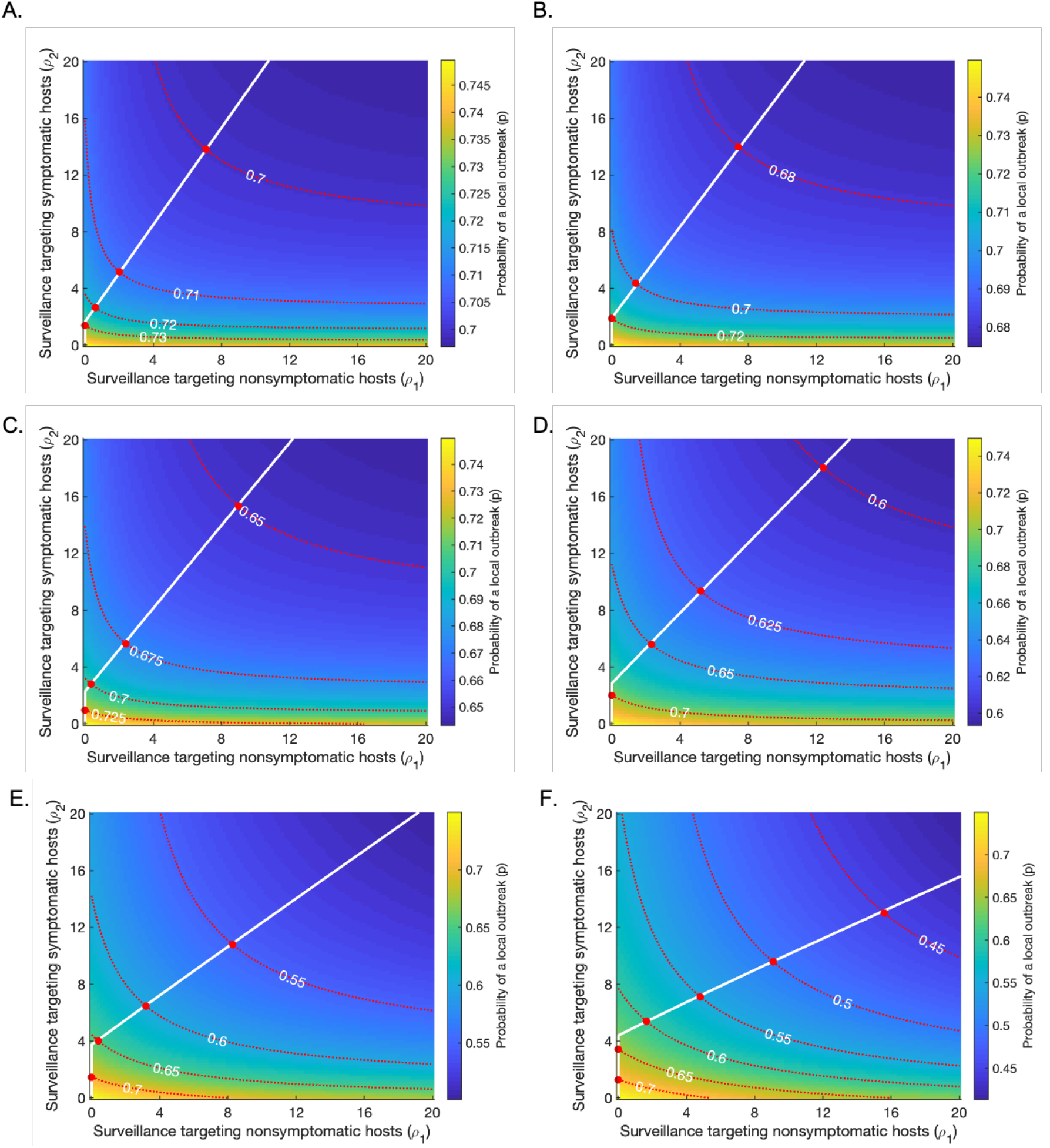
Varying the upper bound on the fractional reduction in the time to isolation (if no other event occurs), *δ*, from its baseline value (*δ* = 0. 8). Plots are analogous to Fig 3D in the main text, showing strategies for minimising the surveillance effort required to achieve a pre-specified risk level (an “acceptable” local outbreak probability). Red dotted lines represent contours along which the probability of a local outbreak is constant, as labelled; red circles indicate the points along these contours at which the total surveillance effort *ρ*_1_ + *ρ*_2_ is minimised. The white line indicates the optimal strategy to follow if the pre-specified risk level is reduced. Apart from *δ*, all parameters are held fixed at their baseline values given in Table 1. A. *δ* = 0.5. B. *δ* = 0.6. C. *δ* = 0.7. D. *δ* = 0.8 (baseline). E. *δ* = 0.9. F. *δ* = 0.95

